# Analysis of SARS-CoV-2 variants from 24,181 patients exemplifies the role of globalisation and zoonosis in pandemics

**DOI:** 10.1101/2021.09.10.21262922

**Authors:** Philippe Colson, Pierre-Edouard Fournier, Hervé Chaudet, Jérémy Delerce, Audrey Giraud-Gatineau, Linda Houhamdi, Claudia Andrieu, Ludivine Brechard, Marielle Bedotto, Elsa Prudent, Céline Gazin, Mamadou Beye, Emilie Burel, Pierre Dudouet, Hervé Tissot-Dupont, Philippe Gautret, Jean-Christophe Lagier, Matthieu Million, Philippe Brouqui, Philippe Parola, Michel Drancourt, Bernard La Scola, Anthony Levasseur, Didier Raoult

**Author notes:** Contributed equally (Philippe Colson, Pierre-Edouard Fournier). Corresponding author: Didier Raoult, IHU Méditerranée Infection, 19-21 Boulevard Jean Moulin, 13005 Marseille, France.

## Abstract

After the end of the first epidemic episode of SARS-CoV-2 infections, as cases began to rise again during the summer of 2020, we at IHU Méditerranée Infection in Marseille, France, intensified the genomic surveillance of SARS-CoV-2, and described the first viral variants. In this study, we compared the incidence curves of SARS-CoV-2-associated deaths in different countries and reported the classification of SARS-CoV-2 variants detected in our institute, as well as the kinetics and sources of the infections. We used mortality collected from a COVID-19 data repository for 221 countries. Viral variants were defined based on ≥5 hallmark mutations shared by ≥30 genomes. SARS-CoV-2 genotype was determined for 24,181 patients using next-generation genome and gene sequencing (in 47% and 11% of cases, respectively) or variant-specific qPCR (in 42% of cases). Sixteen variants were identified by analysing viral genomes from 9,788 SARS-CoV-2-diagnosed patients. Our data show that since the first SARS-CoV-2 epidemic episode in Marseille, importation through travel from abroad was documented for seven of the new variants. In addition, for the B.1.160 variant of Pangolin classification (a.k.a. Marseille-4), we suspect transmission from mink farms. In conclusion, we observed that the successive epidemic peaks of SARS-CoV-2 infections are not linked to rebounds of viral genotypes that are already present but to newly-introduced variants. We thus suggest that border control is the best mean of combating this type of introduction, and that intensive control of mink farms is also necessary to prevent the emergence of new variants generated in this animal reservoir.

## INTRODUCTION

Following its emergence in December 2019 in Wuhan, China, SARS-CoV-2 was declared to be a pandemic in March 2020 and its incidence since then has shown different epidemic patterns according to the country (https://ourworldindata.org/; https://www.ecdc.europa.eu/en/geographical-distribution-2019-ncov-cases) (*1*). In Asia, a bell-shaped curve typical of seasonal viral respiratory infections has been observed. In Western developed countries, successive phases have occurred that have associated an initial bell-shaped curve and additional waves with one or more peaks. In Marseille, France, after a first epidemic period characterised by a bell-shaped curve of incidence followed by a period of almost zero incidence in May and June 2020, an epidemic burst of SARS-CoV-2 cases occurred in July. As this differed from the single bell-shaped curve typically observed for most respiratory viral infections, we suspected this corresponded to the introduction of new SARS-CoV-2 genotypes. Consequently, in the summer of 2020 we expanded our assessment of viral genetic diversity using genome next-generation sequencing. This allowed us to detect and report as early as 7 September 2020 several SARS-CoV-2 lineages that occurred concurrently or successively, and were associated with different genetic, epidemiological and clinical features (*2–4*).

These observations required defining and naming these different lineages to communicate about them, which is the very principle of taxonomy. Classifying and naming emerging infectious agents is essential (*5*) but the task is particularly difficult in the case of SARS-CoV-2 for several reasons. First, as for other RNA viruses, SARS-CoV-2 evolves through continuous genetic variation with a high mutation rate (*6, 7*). Second, SARS-CoV-2 has caused a dramatically high number of infections (over 195 million human cases worldwide and 5.9 million cases in France by 31 July 2021 (https://www.ecdc.europa.eu/en/geographical-distribution-2019-ncov-cases)). In addition, there are animal reservoirs, a major one being mink (*8, 9*). SARS-CoV-2 is, therefore, currently a rapidly evolving virus with a mutation rate estimated to be 9.8 x 10^-4^ substitutions/site/year (*10*) and characterised by a high rate of lineage turnover (*11*). Several SARS-CoV-2 specific classification and naming systems including those of GISAID (https://www.gisaid.org/ (*12*)), Nexstrain (https://clades.nextstrain.org/ (*13*)), Pangolin (https://cov-lineages.org/pangolin.html (*11*)), or World Health Organization (WHO) (https://www.who.int/en/activities/tracking-SARS-CoV-2-variants/). But, although these are of great interest and usefulness, other classification and naming systems are also valuable, as previously acknowledged (*11*), particularly when it comes to better exploring genomic epidemiology on a local scale through real-time next-generation sequencing. Indeed, some classifications have become complicated with the advent of multiple new variants. In addition, local genetic trends may be underrepresented in the tremendous flow of available genomes. Moreover, correlations between genotypic patterns and epidemiological and clinical characteristics can be accurately performed at institutional or local levels through a large network of people who are responsible for patient diagnosis and clinical management, while remote collection and analysis of data may have poorer performance. Thus, we aimed at using our own definition, classification and naming of SARS-CoV-2 lineages and at correlating these lineages with our local epidemiological data in order to better investigate and monitor their genetic features and their sources and geographical origins. Here, we describe our classification and naming system of SARS-CoV-2 lineages and the history and characteristics of the major lineages detected among patients diagnosed at our institute, which sharply highlights the role of travel and globalisation in the current pandemic.

## RESULTS

### Broad diversity according to countries of the patterns of incidence of SARS-CoV-2-associated deaths

Clustering of the incidence curves of SARS-CoV-2-associated deaths for 221 countries delineated ten major groups of countries (Fig. 1A). Each group comprised a mean±standard deviation of 12±4 countries (range, 7–21). A broad range of patterns was revealed through these groups, with countries experiencing one or more epidemic waves of varying intensity and duration combined or not with periods of lower and uneven incidence (Fig. 1B). These data indicate that the incidence of SARS-CoV-2-associated deaths did not follow a single bell-shaped curve except in countries from Group 1, which includes China, Australia and a few countries in Central Asia and the Middle East. Instead, the incidence of SARS-CoV-2-associated deaths evolved elsewhere with multiple epidemic patterns according to time and geographic location. The worldwide distribution of these patterns showed that the majority of Western and Northern European countries (including France), the USA and Canada, which are part of Group 10, shared a same pattern (Figs. 1A and 1C). Similarly, several countries in Asia and South America shared the same patterns leading to their classification in Groups 1–4. In contrast, a broad panel of patterns was observed for Africa. Countries that experienced a single peak of SARS-CoV-2-associated deaths (Groups 1, 4 and 7) were mostly in Asia, Oceania, the Middle East, South America, Africa, and Eastern Europe; exceptions were Luxembourg, Austria and Switzerland. Among the countries which experienced several peaks of SARS-CoV-2-associated deaths, those in Group 10 were the developed industrialised countries of Western Europe and North America. These countries are particularly involved in globalisation through tourism, business, and immigration (*14*). In Marseille and its geographical area, the overall pattern of incidence of SARS-CoV-2-associated deaths was similar to that of countries in Group 10, which includes France, with a bell-shaped curve in March–April 2020 then a short period with a very low incidence followed by a longer period with two peaks of incidence and erratic incidence between these peaks (Fig. 2A). This pattern of incidence with several peaks suggests multiple introductions of new variants that evolved abroad and caused distinct epidemics.

**Fig. 1.**
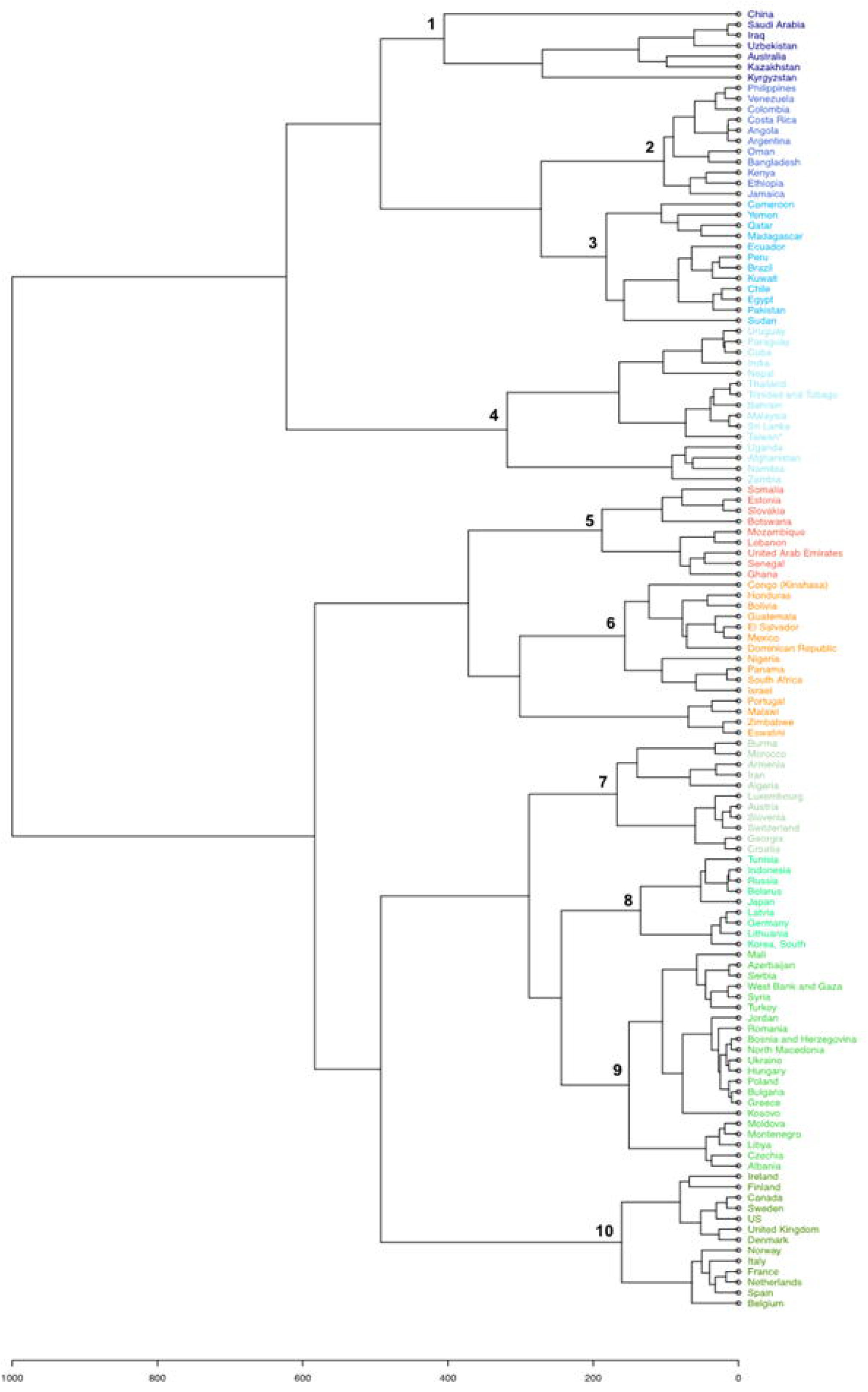
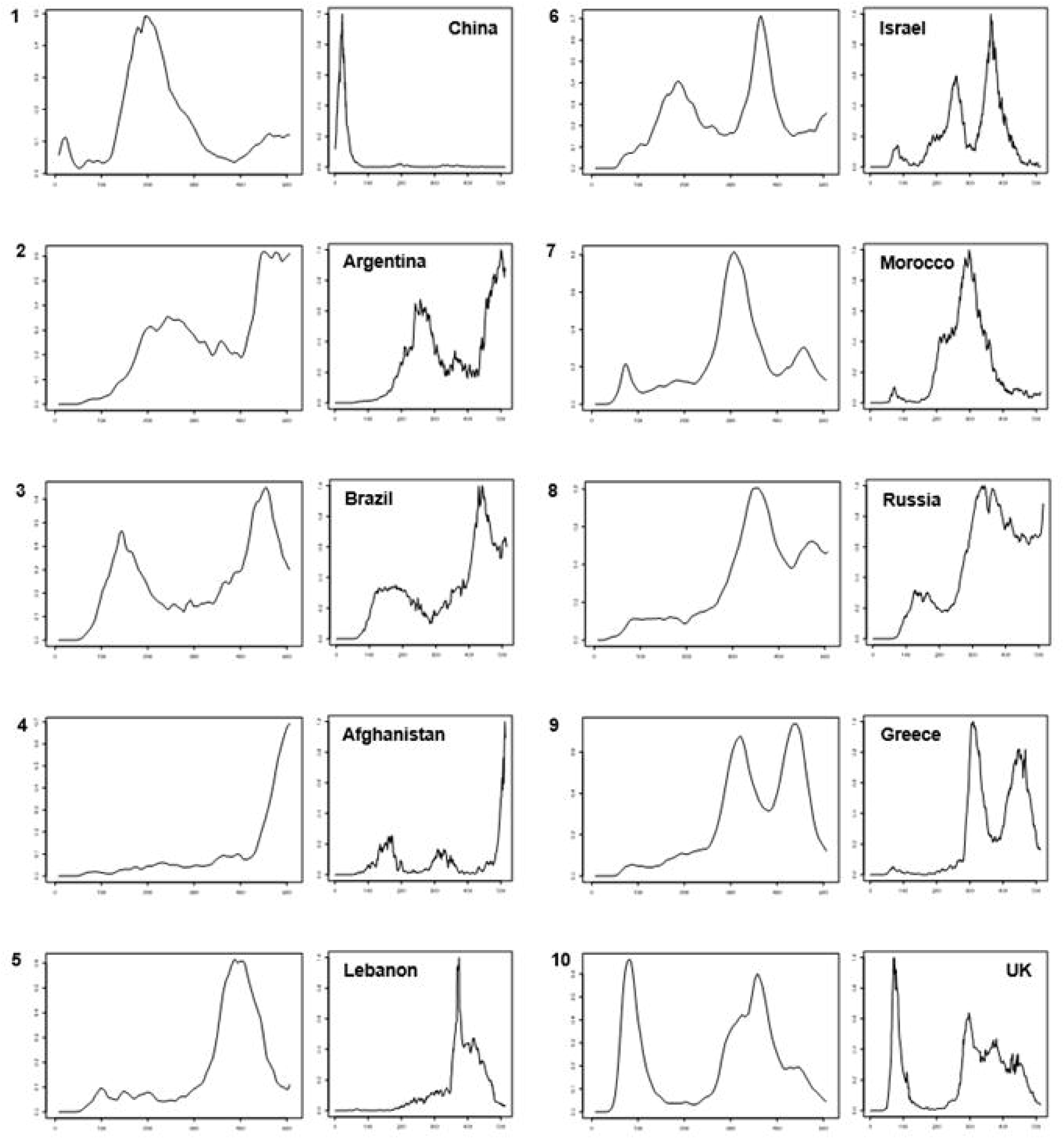
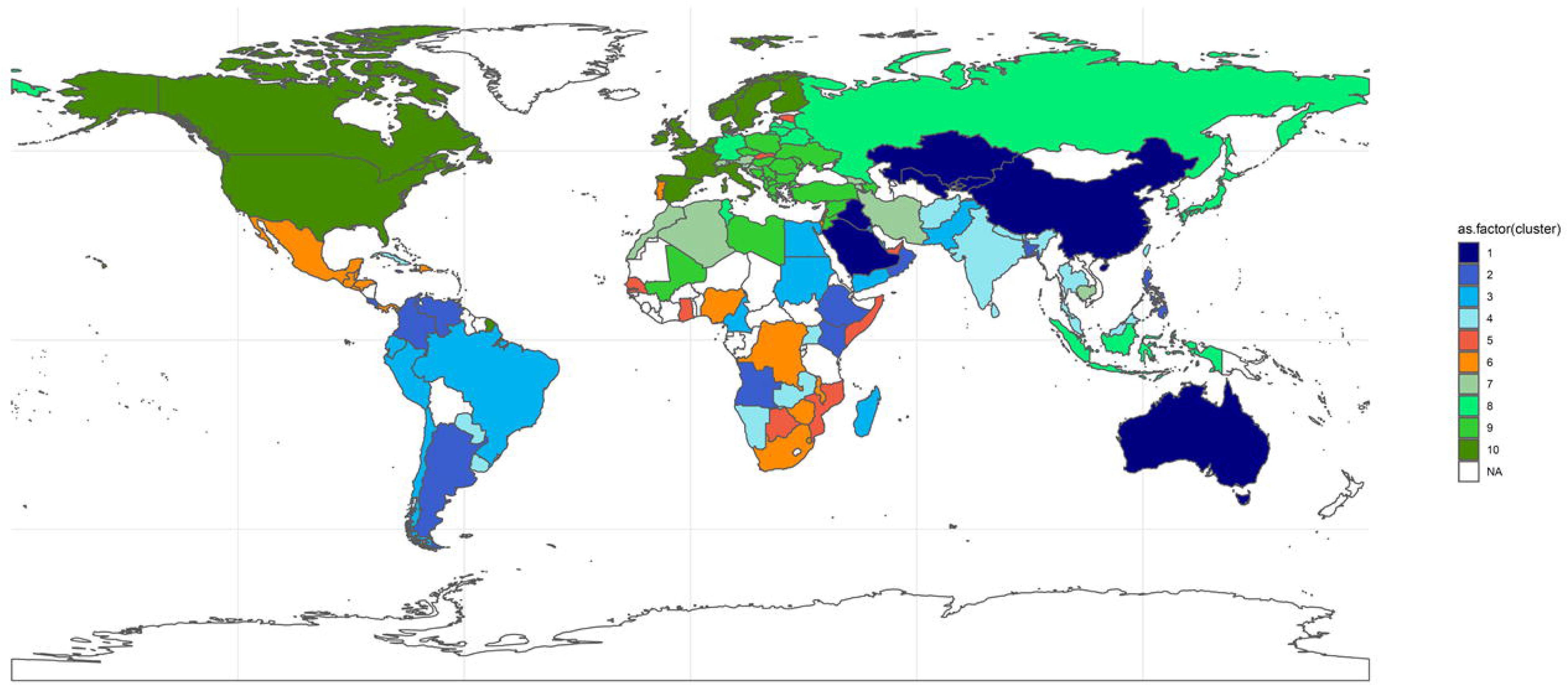
Patterns of incidence curves of SARS-CoV-2-associated deaths according to countries. **(A)** Hierarchical clustering of incidence curves of SARS-CoV-2-associated deaths per country **(B)** Major patterns of incidence curves of SARS-CoV-2-associated deaths defined based on hierarchical clustering, and the example of one country for each pattern. Patterns of incidence curves of SARS-CoV-2-associated deaths are shown in 10 panels numbered from 1 to 10; these numbers correspond to those from the cladogram of Figure 1A. The example of one country is shown in a panel at the right of each of the ten patterns. **(C)** Worldwide distribution of the ten major patterns of incidence curves of SARS-CoV-2-associated deaths defined based on hierarchical clustering. Colours are as used for Figure 1A.

**Fig. 2.**
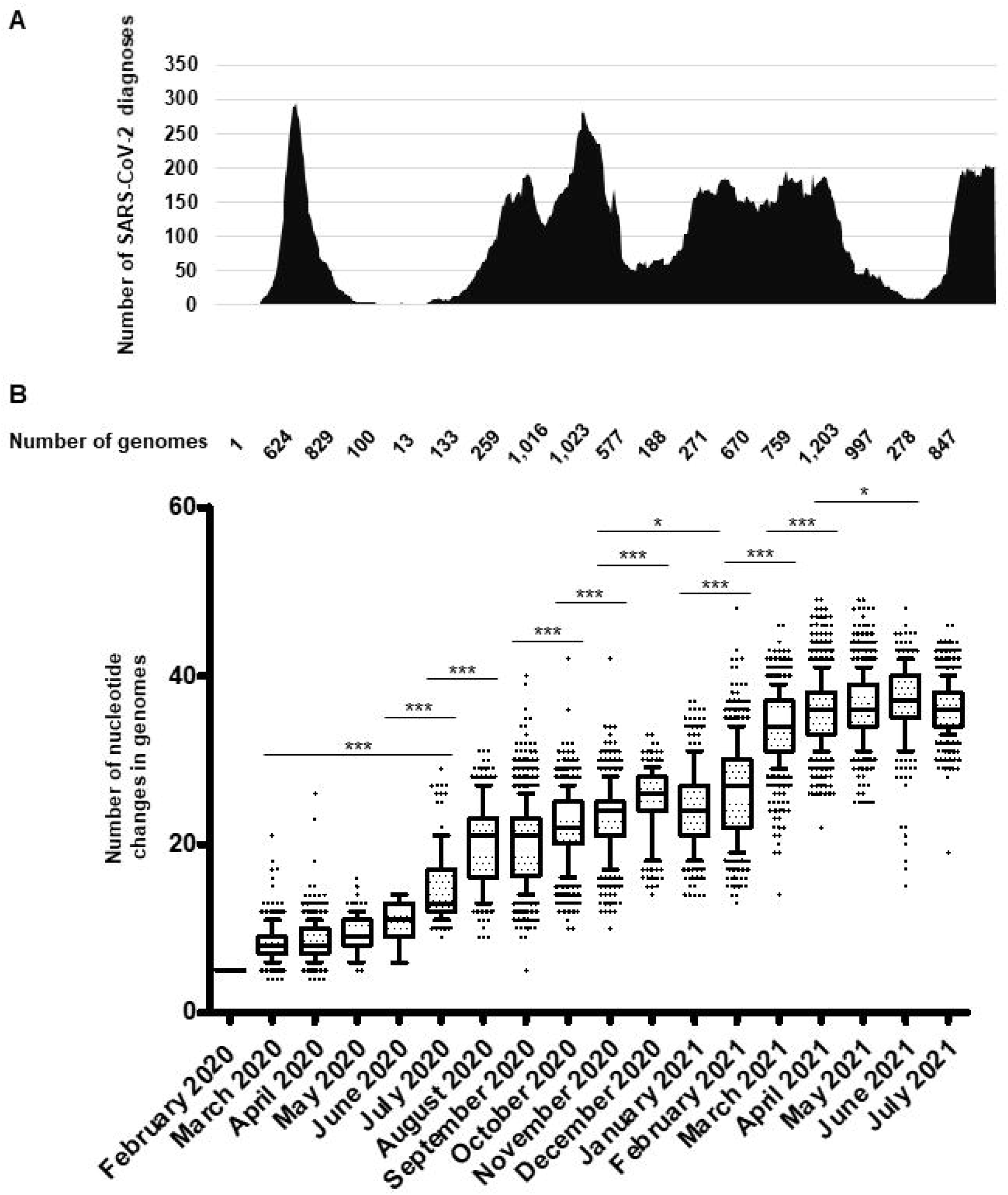
Chronological distribution of SARS-CoV-2 diagnoses by qPCR at IHU Méditerranée Infection institute (A) and of mean (±standard deviation) numbers of mutations in SARS-CoV-2 genomes obtained per month from patients SARS-CoV-2-diagnosed at IHU Méditerranée Infection, Marseille (B). (A) Chronological distribution of SARS-CoV-2 diagnoses is the mean number of diagnoses by qPCR per sliding windows of 7 days and steps of one day. (B) Numbers of mutations in SARS-CoV-2 genomes are calculated in reference to the genome of the Wuhan-Hu-1 isolate (GenBank Accession no. NC_045512.2). Whiskers indicate 10-90 percentiles. ***: <10^-3^; *: p<10^-1^.

### Increase of SARS-CoV-2 genetic diversity over time

A total of 54,703 (10.6%) out of 513,805 patients tested in our institute for SARS-CoV-2 infection between 29 January 2020 and 18 August 2021 were positive by real-time reverse transcription-PCR (qPCR) (https://www.mediterranee-infection.com/covid-19/) (Fig. 2A). We observed a bell-shaped curve of incidence between late February and early May 2020. This was followed by a period with almost no diagnoses (between 0 and 8 per day; mean±standard deviation, 2.5±2.2 per day) which lasted from 9 May to 5 July 2020. We performed whole SARS-CoV-2 genome sequencing using next-generation sequencing procedures, since we diagnosed the first infection on 27 February 2020 (*15*) and primarily obtained viral genomes from 309 patients for the March–April period (*2, 16*). In July 2020, the incidence of SARS-CoV-2 diagnoses increased again (Fig. 2A). As the occurrence of a second epidemic two months after the end of the first one was unexpected for a respiratory virus infection, we suspected that it was linked to the introduction of new SARS-CoV-2 genotypes. This led us to quicky expand the next-generation sequencing of SARS-CoV-2 genomes to test this hypothesis. We primarily obtained 382 additional genomes during summer 2020 (*2*), and ultimately analysed the viral genomes obtained from a total of 9,788 patients sampled between 27 February 2020 and 18 July 2021. We observed a significant increase in the mean number of mutations in SARS-CoV-2 genomes between April 2020 and July 2020 (mean (±standard deviation) number of mutations per genome: 8.5±2.0 versus 15.0±4.4, respectively; p< 10^-3^ (ANOVA test)) (Fig. 2B; Table S1). This mean number continued to increase during the summer of 2020, and the increase was significant between July and August (20.0±5.0). Thereafter, significant increases in the mean number of mutations per genome were observed between September and December (25.2±4.3), between November 2020 (23.0±4.1) and February 2021 (26.4±5.7), and between February and March 2021 (34.1±4.2). Similar trends were observed for the number of amino acid changes in the spike protein (Fig. S1). Overall, when analysing the pairwise genetic distance between genomes obtained over time, two major periods with increased viral genetic diversity compared to previous months were identified, during the summer of 2020 compared to before June 2020, mean±standard deviation of pairwise genetic distances between genomes being 8.28×10^- 4^±3.50×10^-4^ versus 2.34×10^-4^±1.24×10^-4^, respectively (p< 10^-3^), then in January–March 2021 when the mean±standard deviation pairwise genetic distances between genomes was 13.93×10^-4^±4.48×10^-4^ compared to 7.14×10^-4^±4.44×10^-4^ (p< 10^-3^) (Fig. 3).

**Fig. 3.**
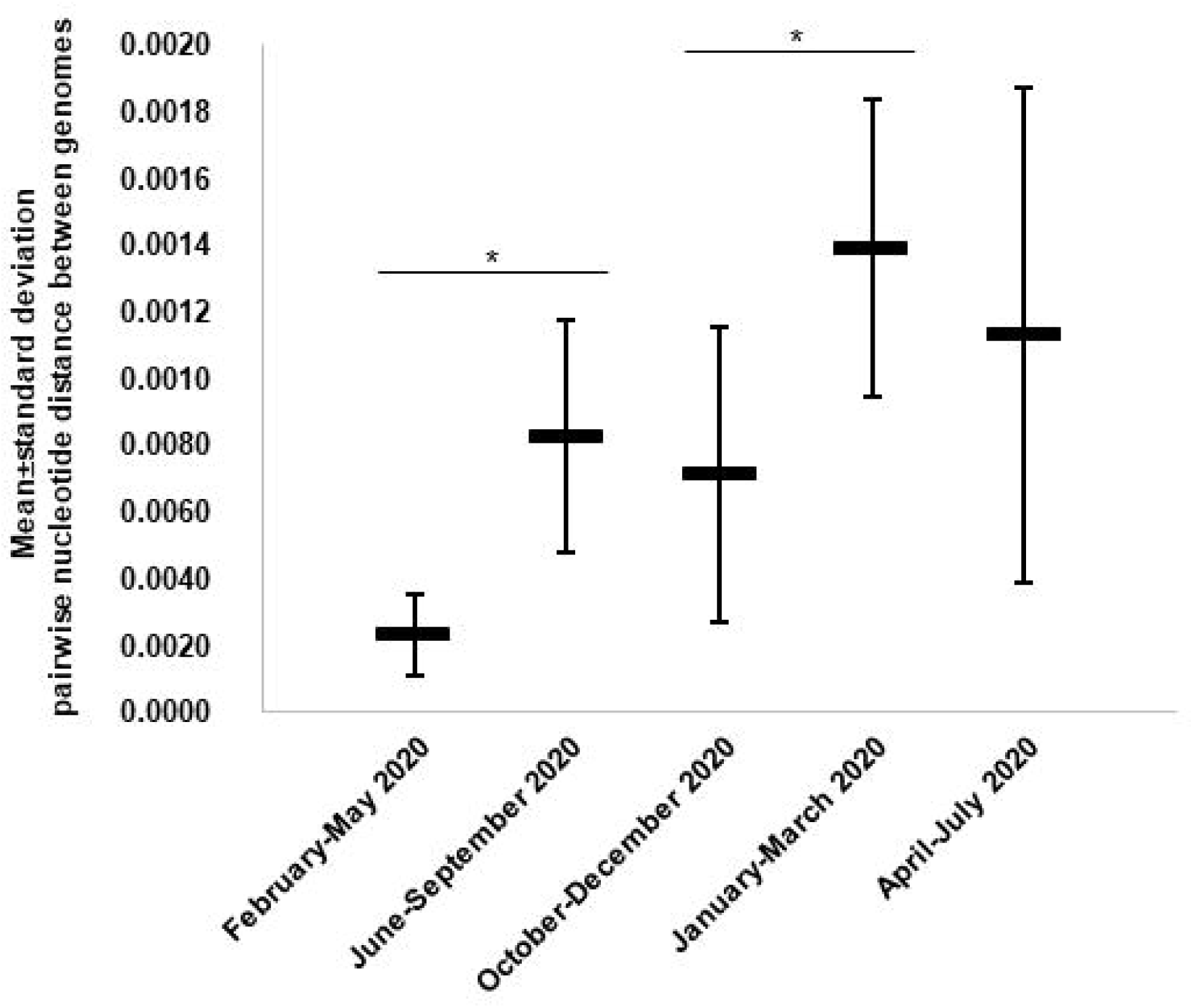
Mean (±standard deviation) pairwise genetic distance between SARS-CoV-2 genomes obtained from patients SARS-CoV-2-diagnosed at IHU Méditerranée Infection, Marseille, according to time periods.

### The Marseille IHU Méditerranée Infection classification system of SARS-CoV-2 variants

Since the summer of 2020, we intensified our activity of SARS-CoV-2 genotyping using next-generation sequencing and also through performing partial spike gene sequencing and implementing variant-specific qPCR (*17, 18*). To characterise the increasing SARS-CoV-2 genome diversity observed during the summer of 2020, we promptly implemented a viral genome classification system and a nomenclature of SARS-CoV-2 variants. The term “SARS-CoV-2 variant” had scarcely been used in the scientific literature before September 2020 (Fig. S2) and no definition had been proposed to differentiate variants from mutants. We first reported the emergence of SARS-CoV-2 variants on 7 September, 2020 (*2*), which included Marseille-2 (later named B.1.177 according to Pangolin classification) and Marseille-4 (B.1.160) variants that were then reported in October 2020 to have emerged and, for the case of Marseille-2, to have spread across Europe (*19*).

There are no universally recognised or used strategies and criteria to classify viruses below the species level, and only biological classifications and nomenclatures have been proposed (*20, 21*) (Box 1). Classification and naming systems have been specifically proposed for SARS-CoV-2 including those of GISAID (https://www.gisaid.org/) (*12*), Nextstrain (https://clades.nextstrain.org/) (*13*), Pangolin (https://cov-lineages.org/pangolin.html) (*11*), and the WHO (https://www.who.int/en/activities/tracking-SARS-CoV-2-variants/) (Box 2). However, we chose to implement our own classification system and nomenclature and to use names rather than only numbers. This approach fitted the viral genomic epidemiology in our geographical area, and was more understandable and easier to use, which is the very purpose of classification and nomenclature. Our strategy consisted in detecting all mutations in all genomes obtained from patients diagnosed in our institute and in assigning a dynamic nomenclature to track SARS-CoV-2 genetic patterns.

#### Box 1. Definition of virus groups at and below species level

**Viral species:** Defined according to a polythetic classification as monophyletic groups of viruses whose properties can be distinguished from those of other species by multiple criteria including increasingly genetic criteria (https://talk.ictvonline.org/taxonomy/w/ictv-taxonomy) (*47*); (*48*).

Below the level of species, there are currently no universally recognised or used strategies and criteria to classify viruses (*20, 21*).

**Viral clade:** Monophyletic group comprised by a common ancestor and all its descent. They are branches with an independent root that obey a Darwinian evolutionary process.

**Viral isolate:** Viral population obtained by culture, generally linked to a particular virus sample (*48*).

**Viral strain:** Virus of a given species with stable and heritable biological, serological, and/or molecular characteristics, and/or links to particular hosts, vectors, or symptoms (*48, 49*).

**Viral quasi-species:** The mutant clouds (or swarms) with closely related genomes generated according to a Darwinian evolutionary process. It includes the accumulation of mutations during the replication of RNA viruses through continuous genetic variation, with possible genetic rearrangement by genetic recombination or genome segment reassortment, followed by competition among the mutants generated and selected in a given environment (*7, 50–52*). **Viral mutant:** The result of nucleotide substitutions (non-synonymous or synonymous), deletions or insertions. These nucleotide changes occur randomly, gradually, and spontaneously, according to the Darwinian model (*6, 53, 54*) and possibly at sites critical for viral protein conformation, antigenicity, susceptibility to immune responses, and functionality, including ability to interact with other proteins and enzymatic properties (*37, 55*).

**Viral variant:** An isolate that differs slightly by its transmission mode or the symptoms it causes from a reference virus (*21, 48*) or whose consensus genome sequence differs from that of a reference virus (*21, 48, 49*). Notwithstanding, no definition of the term “variant” is universal or broadly accepted (*21, 56*).

#### Box 2. Major SARS-CoV-2 classification systems

**World Health Organization (WHO) classification (https://www.who.int/en/activities/tracking-SARS-CoV-2-variants/):**

Defines SARS-CoV-2 ‘Variants of Interest’ (VOI) and ‘Variants of Concern’ (VOC). **VOI:** viral genome has mutations with established or suspected phenotypic implications and either has been identified to cause community transmission/multiple COVID-19 cases/clusters or has been detected in multiple countries. **VOC:** a VOI demonstrated through comparative assessment to be associated with

(ii) increase transmissibility or detrimental change in COVID-19 epidemiology; (ii) increase in virulence or change in clinical disease presentation; or (iii) decrease in effectiveness of public health and social measures or available diagnostics, vaccines, and/or therapeutics. Nomenclature uses letters of the Greek alphabet (currently from Alpha to Theta as of 1 August 2021).

**GISAID classification (https://www.gisaid.org/references/statements-clarifications/clade-and-lineage-nomenclature-aids-in-genomic-epidemiology-of-active-hcov-19-viruses/)** (***12***): Defines high-level phylogenetic clades informed by the statistical distribution of genome distances in phylogenetic clusters followed by merging of smaller lineages into major clades (eight currently) based on shared marker mutations. Nomenclature uses actual letters of marker mutations (S, L, V, G, GH, GR, GV, GRY).

**Nextstrain classification ((https://nextstrain.org/blog/2021-01-06-updated-SARS-CoV-2-clade-naming) (*13***): Defines major clades that reach ≥20% global frequency for ≥two months, ≥30% regional frequency for ≥two months, or correspond to a ‘variant of concern’ (VOC). Nomenclature uses a year-letter naming system.

**Pangolin (for Phylogenetic Assignment of Named Global Outbreak LINeages) classification (https://cov-lineages.org/pangolin.html)** (***11***): Based on a maximum likelihood phylogenetic reconstruction with the following criteria: (i) Phylogenetic evidence of emergence from an ancestral lineage into another geographically distinct population; (ii) ≥one shared nucleotide difference(s) from the ancestral lineage; (iii) ≥five genomes; (iv) ≥one shared nucleotide change(s) among genomes from the lineage; and (v) a bootstrap value >70% for the lineage node. Regarding the nomenclature: a letter (A for the Wuhan-Hu-1 isolate) is given first, followed by numbers for a maximum of three sublevels after which new descendant lineages are given a new letter.

**Marseille classification:** Defines groups of viral genomes carrying specific sets of ≥five mutations differentiating them from any other viral genomes and obtained from ≥30 different patients. Nomenclature uses our city name, “Marseille”, followed by a number according to the chronology of their description (e.g.: Marseille-1, Marseille-2,…) or to one of their hallmark amino acid substitutions (e.g.: Marseille-501).

A phylogenetic tree was built that included all genomic sequences of SARS-CoV-2 obtained in our institute (Fig. 4A). Based on tree topology and to distinguish them from SARS-CoV-2 mutants, we delineated SARS-CoV-2 variants as groups of viral genomes carrying a set of at least five mutations differentiating them from any other viral genomes and obtained from at least 30 (initially 10) different patients. Genomes were, therefore, unambiguously assigned to a given variant based on the presence of particular sets of mutations (Table 1). This differentiated variants from mutants, the genomes of which do not harbour this minimal set of hallmark mutations. We first delineated seven Marseille variants of SARS-CoV-2 as early as 7 September 2020, 2 of which did not eventually reach the threshold number of 30 genomes (*2*). This was the first description of an emergence and expansion of several SARS-CoV-2 variants responsible for distinct epidemics in a same geographical area. Three additional variants were then defined in 2020 (Fig. 5) and four in 2021, making a total of 12 (Table 1; Figs. 4A and 4B). In addition, we delineated 9 other viral lineages comprising genomes harbouring a specific set of at least five mutations but obtained from <30 different patients; some of these lineages may grow and become *bona fide* variants as additional members will be obtained. Besides, we detected four other SARS-CoV-2 variants that were defined as variants of concern (VOC; Box 2) and were named Alpha, Beta, Gamma and Delta by the WHO (https://www.who.int/en/activities/tracking-SARS-CoV-2-variants/). Regarding the nomenclature of the variants we defined, we used our city name, Marseille, followed by a number according to the chronology of their description (e.g.: Marseille-1, Marseille-2,…) or one of their hallmark amino acid substitutions (e.g.: Marseille-501). Hereafter, we will use both the Marseille and Pangolin nomenclatures to describe the genotypes of the SARS-CoV-2 genomes obtained in our institute.

**Fig. 4.**
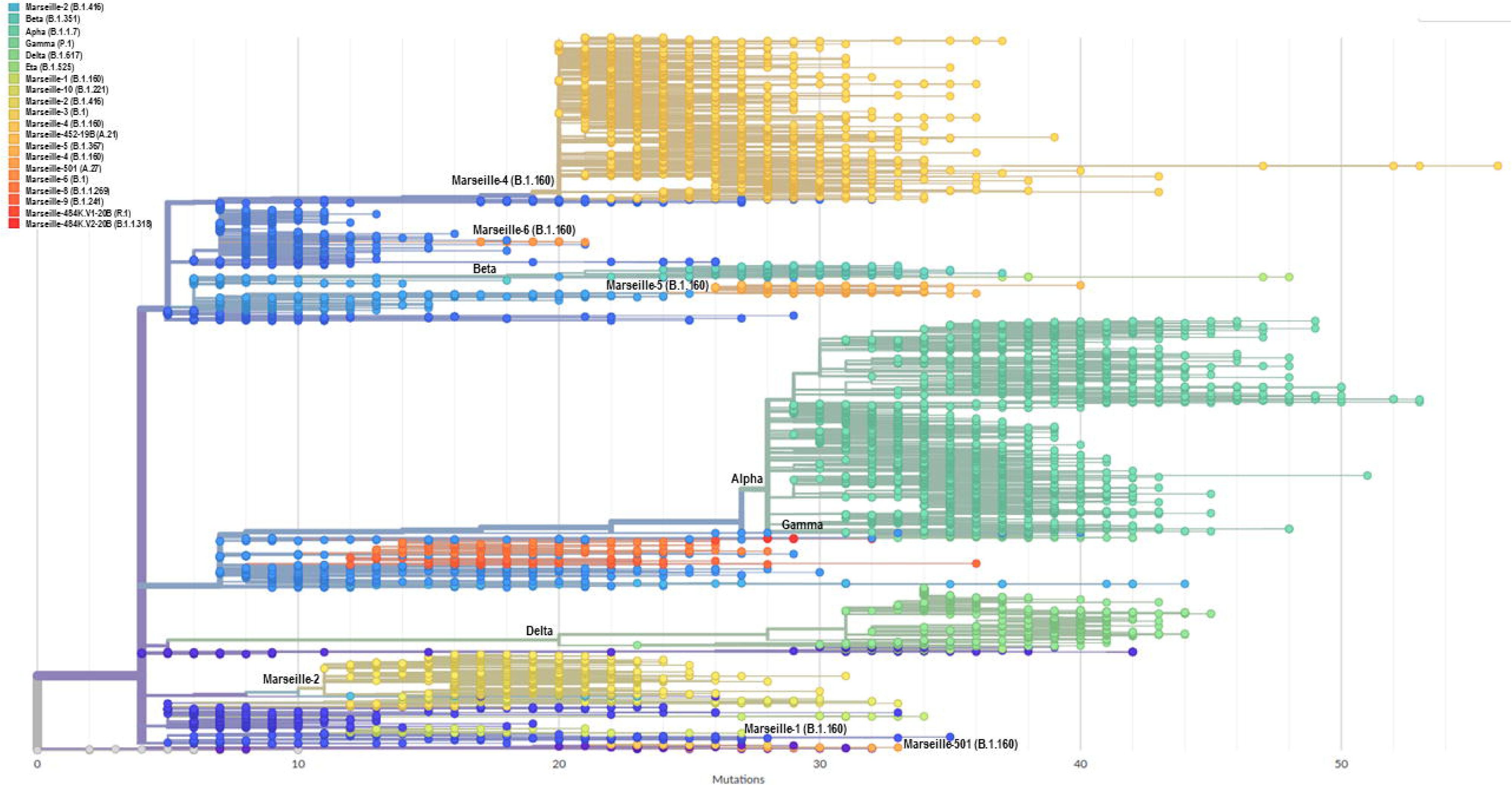
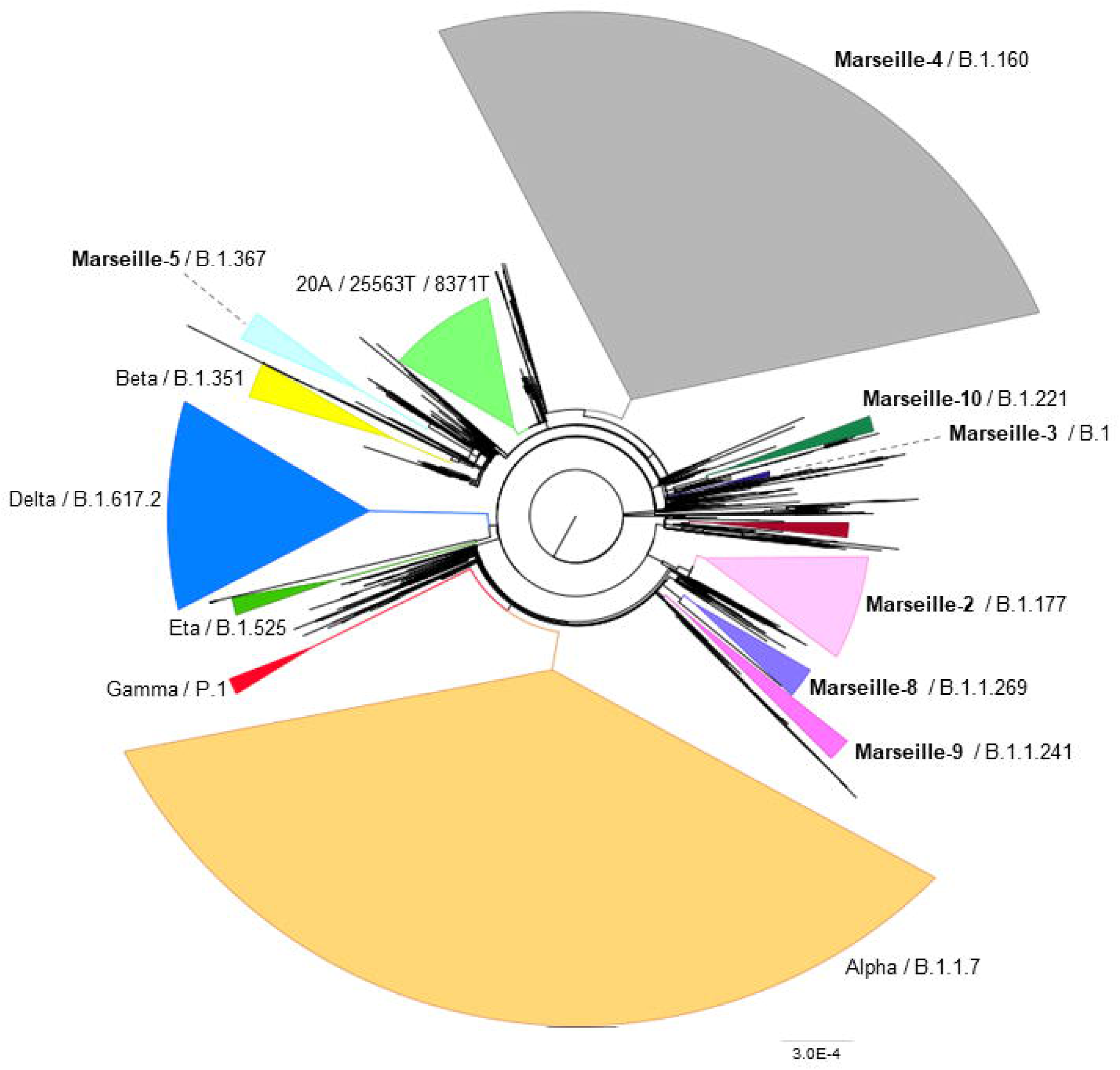
Phylogenetic trees of 10,773 genomic sequences of SARS-CoV-2 obtained from patients SARS-CoV-2-diagnosed at IHU Méditerranée Infection, Marseille. Phylogeny reconstruction was performed using the nextstrain/ncov tool (https://github.com/nextstrain/ncov) then visualised with Auspice (https://docs.nextstrain.org/projects/auspice/en/stable/). The genome of the original Wuhan-Hu-1 coronavirus isolate (GenBank accession no. NC_045512.2) was added as outgroup. Major (most prevalent) variants are labelled. Marseille variants or WHO clades, and Pangolin, or Nextstrain clades are indicated. **(A)** X-axis shows the number of mutations compared to the genome of the Wuhan-Hu-1 isolate (GenBank accession no. NC_045512.2). **(B)** Circular representation of the phylogenetic tree based on 10,773 genomic sequences of SARS-CoV-2 obtained from patients SARS-CoV-2-diagnosed at IHU Méditerranée Infection, Marseille.

**Fig. 5.**
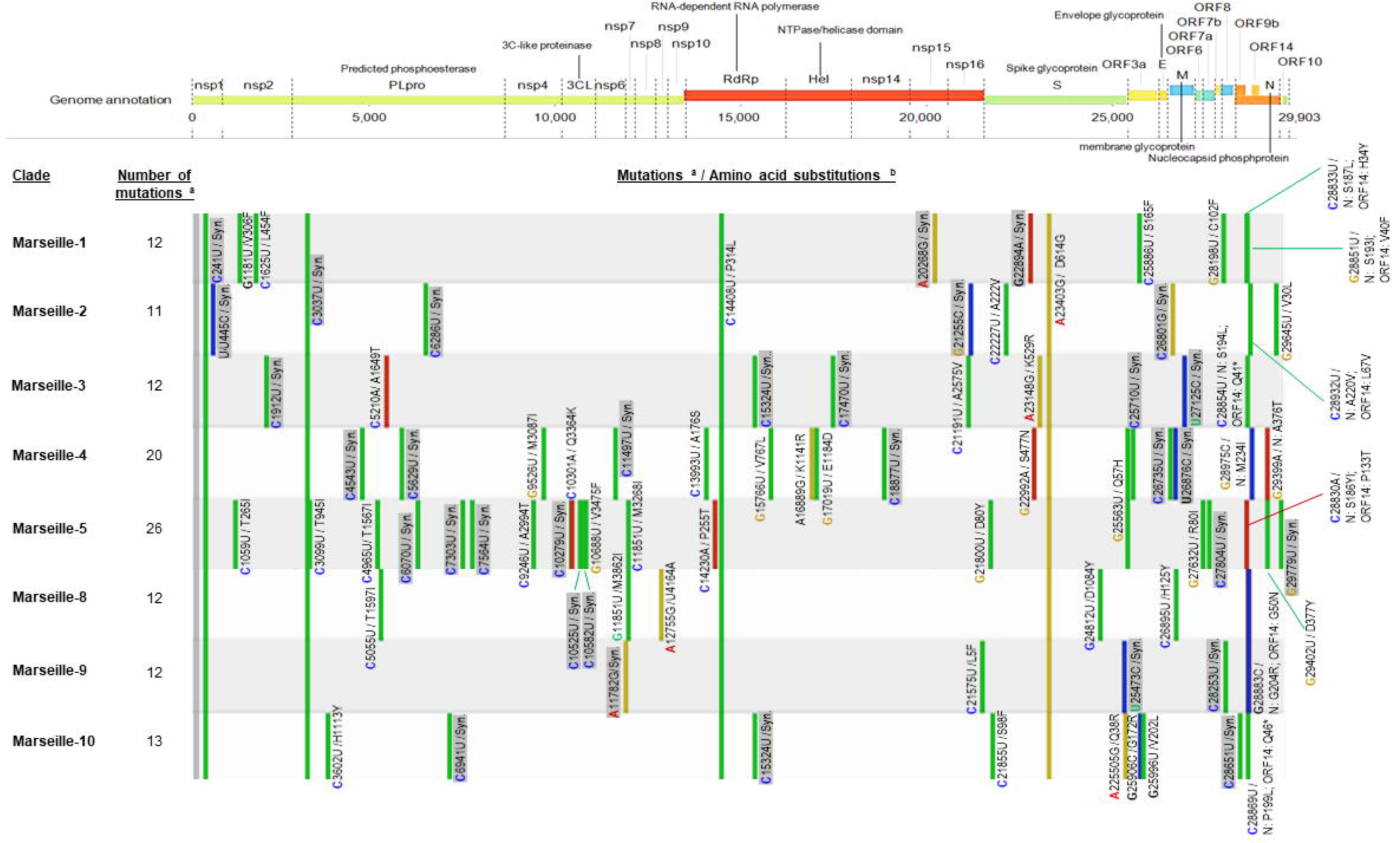
Microarray showing the distribution along the viral genome and in viral genes of nucleotide and amino acid substitutions observed in comparison with the genome of Wuhan-Hu-1 isolate for the various viral variants detected in 2020 in respiratory samples from patients diagnosed with SARS-CoV-2 infection at IHU Méditerranée Infection. Sequences from complete genomes that were obtained were analyzed using the Nextstrain web-tool (https://clades.nextstrain.org/) (*13*). Representation is adapted from Nextclade sequence analysis web application output (https://clades.nextstrain.org/). ^a^ In reference to genome GenBank Accession no. NC_045512.2 (Wuhan-Hu-1 isolate); b Color code for nucleotide mutations: Green: U; yellow: G; blue: C; red: A. nsp9: ssRNA-binding protein; nsp14: 3’-to-5’; exonuclease; nsp15: EndoRNase; PLproSyn.: synonymous.

**Table 1.**
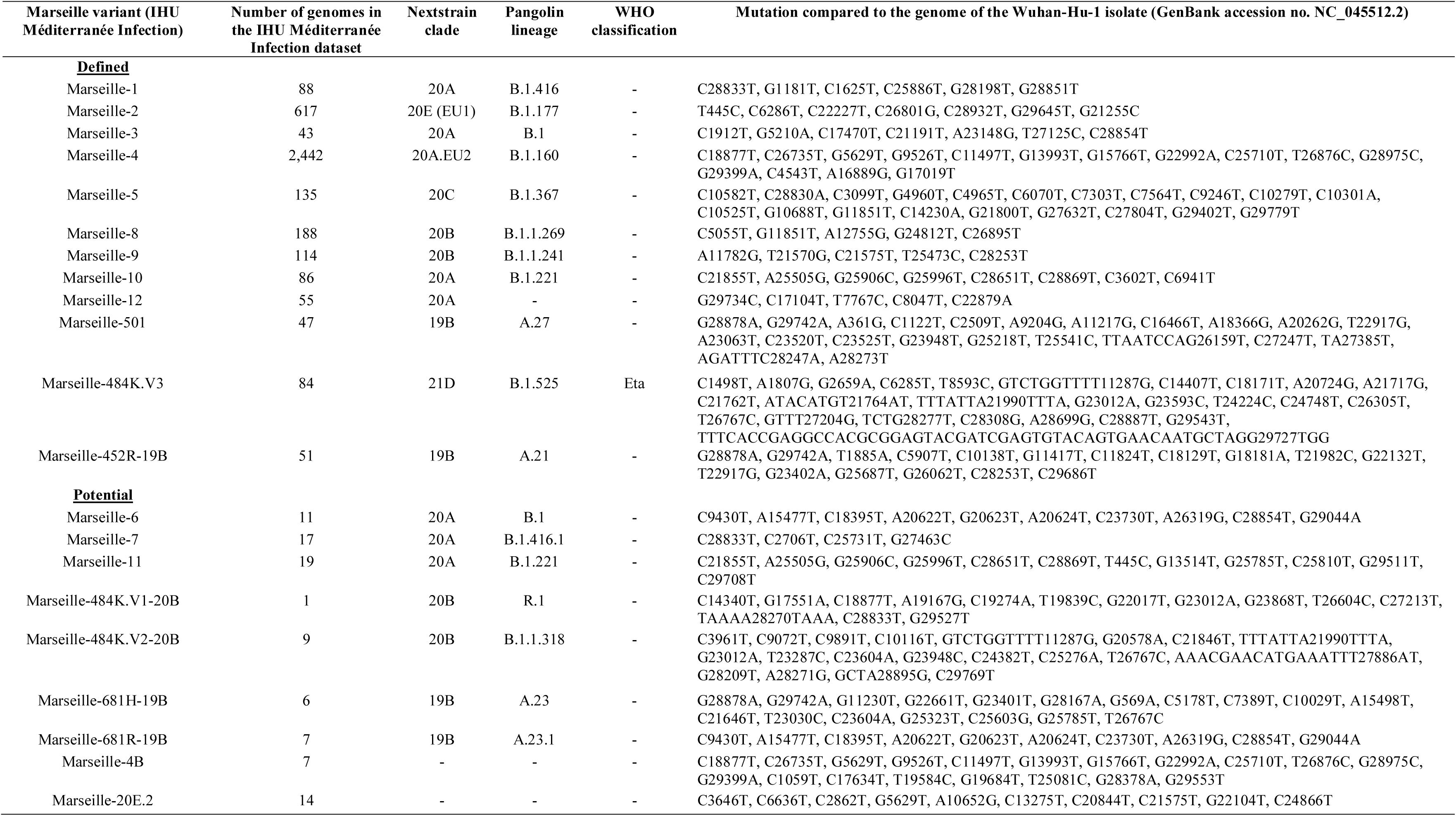
**Nomenclature and description of Marseille (IHU Méditerranée Infection) variants of SARS-CoV-2**

### Emergence and outcome of the SARS-CoV-2 variants that circulated in our geographical area

#### Overall prevalence of the Marseille SARS-CoV-2 variants

SARS-CoV-2 genotyping was performed and successful for 24,181 (44%) of the 54,703 SARS-CoV-2-positive patients diagnosed in our institute as of 18 August 2021 (Table 2). SARS-CoV-2 genotyping was performed by genome sequencing in our institute for at least 12% of positive diagnoses of SARS-CoV-2 infection monthly, and for at least one fifth and one third of positive diagnoses monthly for 14 and 7 of the 18 months from February 2020 to July 2021, respectively (Table S2). Genotype was obtained primarily by whole genome next-generation sequencing in 11,387 (47%) cases and by partial genome next-generation sequencing (spike fragment) in 2,621 (11%) cases, and retrospectively or prospectively (since January 2021) by in house variant-specific qPCR assays gradually designed and implemented subsequent to the detection of new variants (*17, 18*) in 10,173 (42%) cases. The most frequently detected variants were, in decreasing order, the Alpha (a.k.a. UK) variant, first detected in late December 2020 (Pangolin lineage B.1.1.7; n= 9,294 patients (38.4%)); the Marseille-4 variant, first detected in July 2020 (B.1.160; 6,125 (25.3%)); the Delta (a.k.a. Indian) variant, first detected in April 2021 (B.1.617.2; 4,113 (17.0%)); the Marseille-2 variant, first detected in August 2020 (B.1.177; 850 (3.5%)); and the Beta (a.k.a. South African) variant, first detected in January 2020 (B.1.351; 575 (2.4%)). Other variants of significant prevalence and/or interest in our geographical area were the Marseille-8 variant (B.1.1.269; 176 (0.7%)); the Marseille-5 variant (B.1.367; 124 (0.5%)); the Marseille-1 variant (B.1.416; 123 (0.5%)); the Marseille-484K.V3 variant (B.1.525; 112 (0.5%)), the Gamma (a.k.a. Brazilian) variant (P.1; 87 (0.4%)); the Marseille-10 variant (B.1.221; 91 (0.4%)); and the Marseille-501 variant (A.27; 74 (0.3%)).

**Table 2.** Number of the main SARS-CoV-2 variants and mutants diagnosed among the IHU Méditerranée Infection cohort of SARS-CoV-2-infected patients as of 18 August 2021

| Variant/mutant name | Genotyping approach |  |  | Total | % of all genotypes |
| --- | --- | --- | --- | --- | --- |
|  | Next-generation genome sequencing | Next-generation spike gene fragment sequencing | qPCR |  |  |
| Alpha (B.1.1.7, a.k.a. UK) | 3,506 | 2,220 | 3,568 | 9,294 | 38.44 |
| Marseille-4 (B.1.160) | 2,564 | 45 | 3,516 | 6,125 | 25.33 |
| Delta (B.1.617.2, a.k.a. Indian) | 1,406 | 0 | 2,707 | 4,113 | 17.01 |
| Marseille-2 (B.1.177) | 639 | 6 | 205 | 850 | 3.52 |
| Beta (B.1.351, a.k.a. South African) | 204 | 210 | 161 | 575 | 2.38 |
| 20A/8371T (1<sup>st</sup> period) | 488 | 0 | 0 | 488 | 2.02 |
| 20A (1<sup>st</sup> period) | 323 | 44 | 0 | 367 | 1.52 |
| 20C (1<sup>st</sup> period) | 310 | 0 | 0 | 310 | 1.28 |
| 20B (1<sup>st</sup> period) | 290 | 6 | 0 | 296 | 1.22 |
| 20A/15324T (1<sup>st</sup> period) | 176 | 4 | 0 | 180 | 0.74 |
| Marseille-8 (B.1.1.269) | 174 | 2 | 0 | 176 | 0.73 |
| Marseille-5 (B.1.367) | 124 | 0 | 0 | 124 | 0.51 |
| Marseille-1 (B.1.416) | 123 | 0 | 0 | 123 | 0.51 |
| 20A/25563T (1<sup>st</sup> period) | 123 | 0 | 0 | 123 | 0.51 |
| Marseille-9 (B.1.1.241) | 117 | 0 | 0 | 117 | 0.48 |
| Marseille-484K.V3 (B.1.525) | 112 | 0 | 0 | 112 | 0.46 |
| 20A/20268G (1<sup>st</sup> period) | 95 | 0 | 0 | 95 | 0.39 |
| Marseille-10 (B.1.221) | 89 | 2 | 0 | 91 | 0.38 |
| 20A/2416T (1<sup>st</sup> period) | 89 | 0 | 0 | 89 | 0.37 |
| Gamma (P.1, a.k.a. Brazilian) | 70 | 1 | 16 | 87 | 0.36 |
| Marseille-501 (A.27) | 26 | 48 | 0 | 74 | 0.31 |
| 20E (B.1.177) | 56 | 5 | 0 | 61 | 0.25 |
| Marseille-452R-19B (A.21) | 43 | 2 | 0 | 45 | 0.19 |
| Marseille-3 (B.1) | 39 | 0 | 0 | 39 | 0.16 |
| 19B (1<sup>st</sup> period) | 13 | 23 | 0 | 36 | 0.15 |
| Marseille-12 | 36 | 0 | 0 | 36 | 0.15 |
| 20D (B.1.1.1) | 31 | 2 | 0 | 33 | 0.14 |
| Marseille-484K.V2 (B.1.1.318) | 20 | 1 | 0 | 21 | 0.09 |
| Marseille-11 (B.1.221) | 19 | 0 | 0 | 19 | 0.08 |
| 19A (1<sup>st</sup> period) | 15 | 0 | 0 | 15 | 0.06 |
| B.1.214 (a.k.a. Belgian) | 15 | 0 | 0 | 15 | 0.06 |
| Marseille-7 (B.1.416.1) | 15 | 0 | 0 | 15 | 0.06 |
| Marseille-6 (B.1) | 9 | 0 | 0 | 9 | 0.04 |
| Marseille-681R-19B (A.23.1) | 7 | 0 | 0 | 7 | 0.03 |
| Marseille-681H-19B (A.23) | 6 | 0 | 0 | 6 | 0.02 |
| Epsilon (B.1.427/B.1.429) | 6 | 0 | 0 | 6 | 0.02 |
| B.1.621 | 6 | 0 | 0 | 6 | 0.02 |
| Marseille-484.V1 (A.23) | 3 | 0 | 0 | 3 | 0.01 |
| Total | 11,387 | 2,621 | 10,173 | 24,181 | 100.0 |

#### Dynamics of SARS-CoV-2 variants in the IHU Méditerranée Infection cohort of SARS-CoV-2-infected patients

Based on our definition of variants, we were able to identify several since the summer of 2020, which we would currently call variants of concern (VOC) (Box 2). After having defined and named the Marseille variants, we attempted to characterise their dynamic and to analyse their epidemiological features. For some of them, we were able to find their source. During the onset of the SARS-CoV-2 epidemic in late February 2020, we observed two distinct SARS-CoV-2 genotypes that did not make it possible to define variants based on our classification system. They corresponded to Nextstrain clades 19A and 20B that briefly co-circulated in our geographical area during the first period of the SARS-CoV-2 epidemic (Table 2; Figs. 6A, 6B, and 7) (*22, 23*). The first clade was diagnosed from 14 patients in March 2020. The second, which corresponds to viruses harbouring the D614G substitution in the spike, was initially retrieved from patients who stayed in northwest Italy and at the French/Italian border, and quickly established in the Marseille area as a majority clade together with clades 20A and 20C until May 2020.

**Fig. 6.**
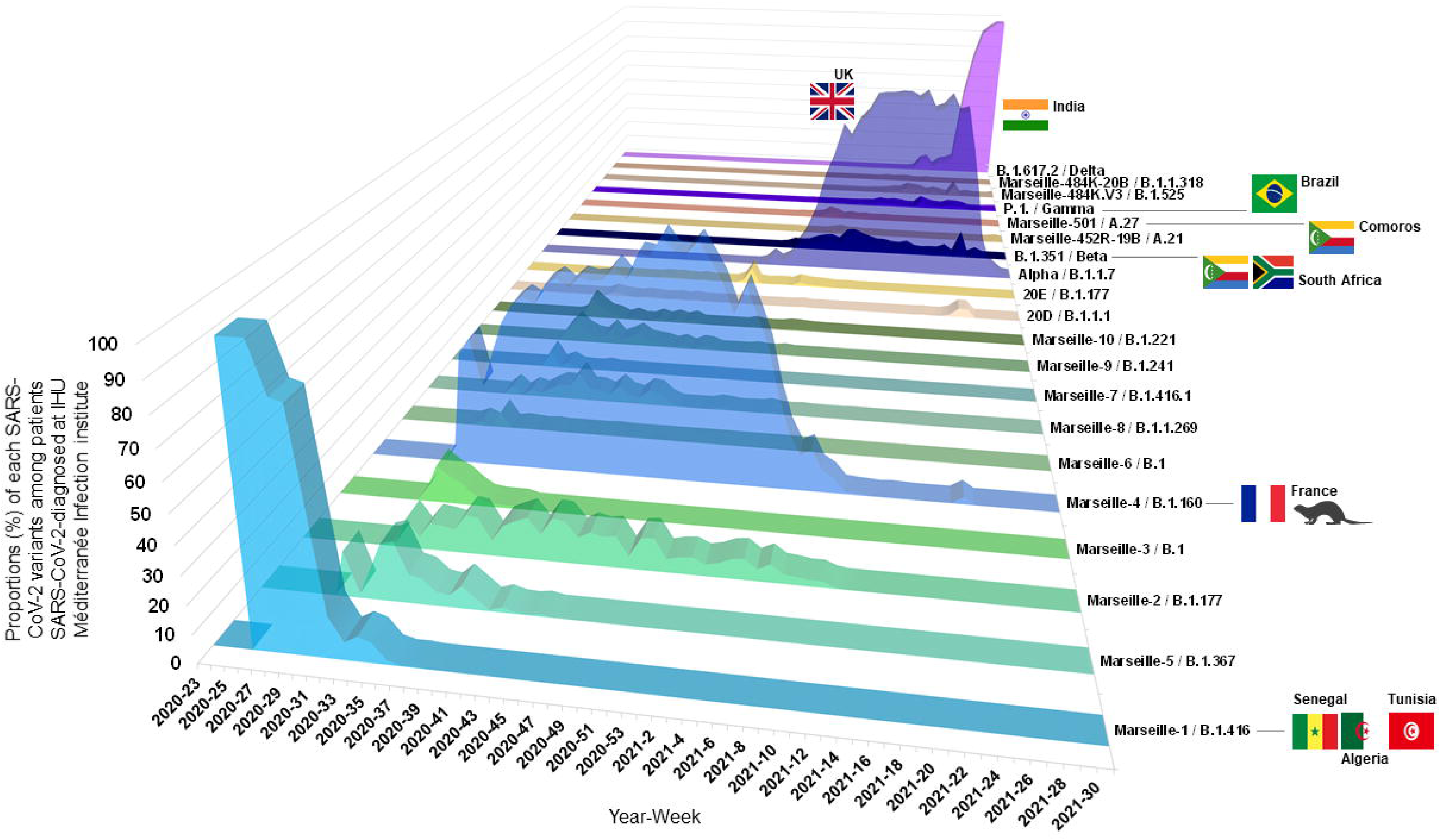
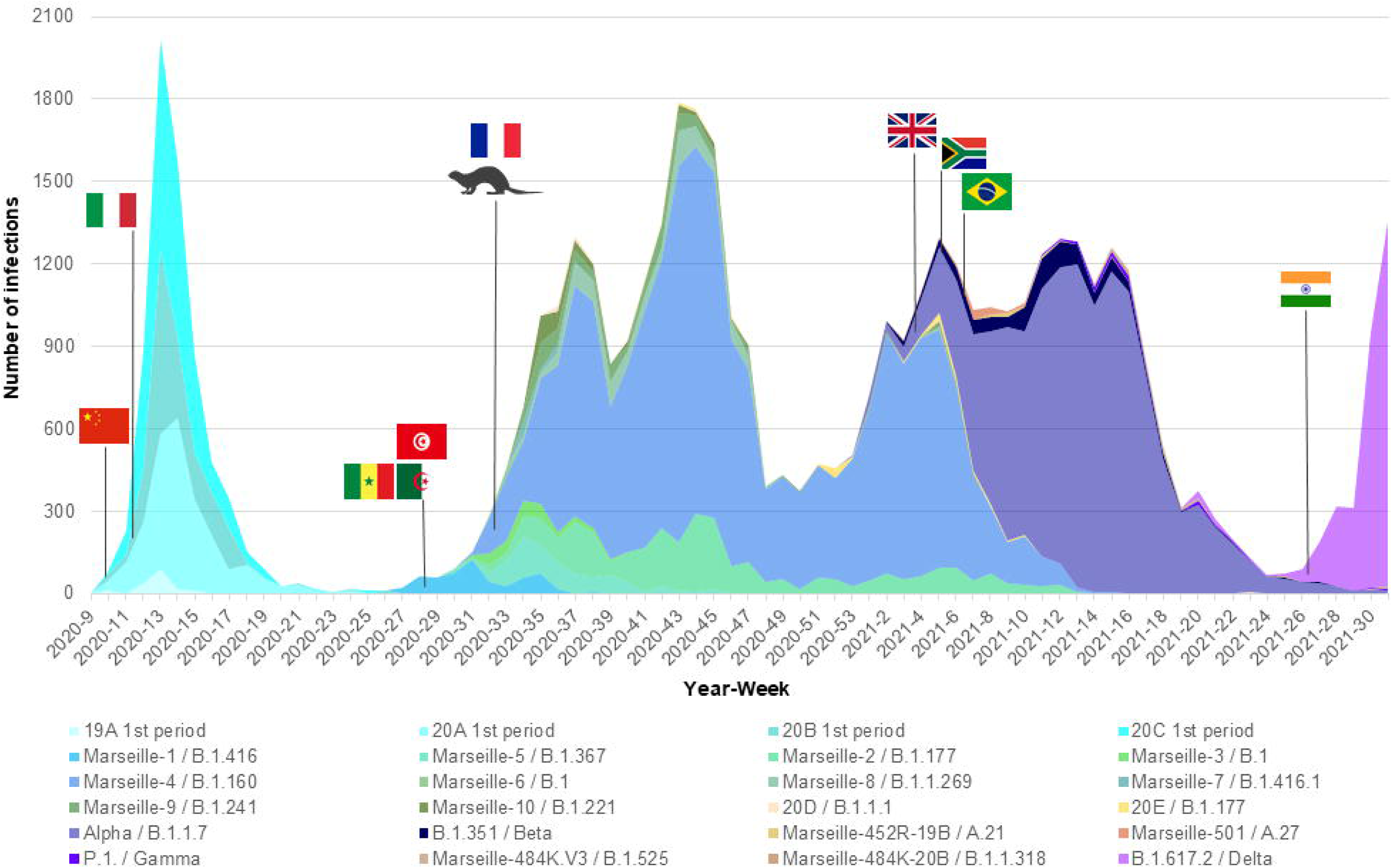
Weekly incidence of each SARS-CoV-2 variants among patients SARS-CoV-2-diagnosed at IHU Méditerranée Infection institute. **(A)** Three-dimensional plot of weekly proportions accounted by each SARS-CoV-2 mutants and variants among patients SARS-CoV-2-diagnosed. **(B)** Weekly incidence of each SARS-CoV-2 mutants and variants extrapolated to the total number of cases, based on their proportions of genotyped cases.

**Fig. 7.**
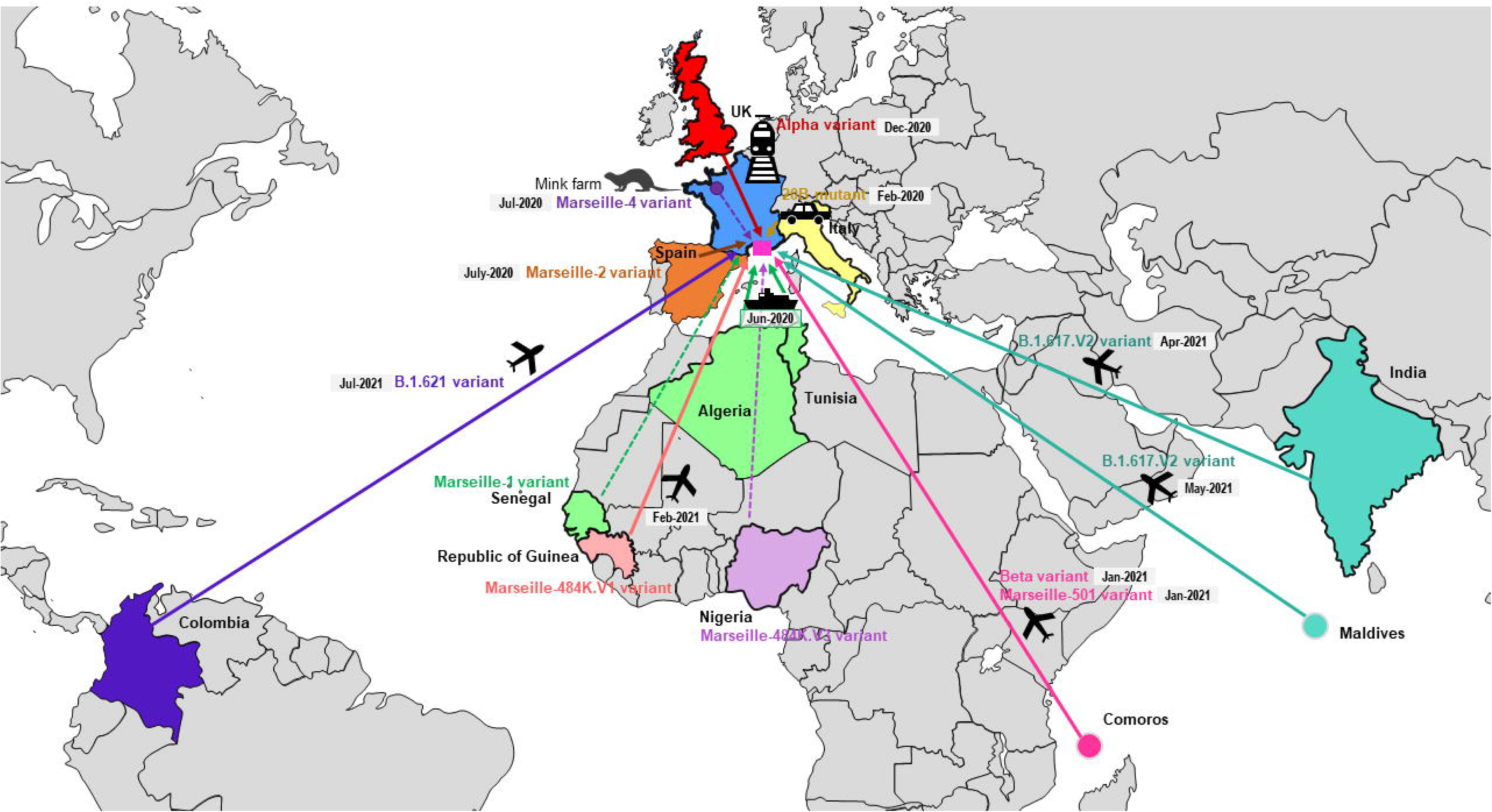
Demonstrated or likely sources and origins of SARS-CoV-2 mutants or variants detected among patients SARS-CoV-2-diagnosed in our institute.

We named the first variant we defined Marseille-1 (corresponds to Pangolin clade B.1.416), which emerged on week 27 of 2020 and predominated in July 2020 before rapidly disappearing at the end of August 2020 (*3*) (Table S3; Figs. 6A, 6B, and 7) (*22, 23*). Its onset coincided with the resumption on 1 July 2020 of maritime connections between Marseille and the Maghreb. As 16 of the first 20 diagnosed cases (80%) were detected in people who had travelled by boat or on passenger boats circulating between the Maghreb and Marseille, we assumed that this variant was brought to Marseille from the Maghreb. Furthermore, genomic analyses showed that it originated from sub-Saharan Africa, mostly Senegal (*3*).

The Marseille-4 variant (B.1.160) was the most prevalent SARS-CoV-2 variant in our geographical area in 2020 and was only outdone by the Alpha (UK) variant in 2021 (*4*). It replaced the Marseille-1 variant in August 2020 and faded away in April 2021. It was first detected in south-west France and in Marseille. Its emergence coincided with the re-increase in France of SARS-CoV-2 incidence starting with a department in northwest France (Mayenne) close to the Eure-et-Loir department where a mink farm was found to be massively infected with SARS-CoV-2 (*4, 24*). The only genome from this farm, obtained from a mink sampled in mid-November 2020 but released in March 2021, is a Marseille-4 variant (GISAID Accession no. EPI_ISL_1392906). In addition, the co-occurrence of 13 hallmark mutations in the Marseille-4 variant without any available genome showing a gradual appearance of these mutations suggested that genetic evolution had been overlooked. These data led us to conclude, based on genomic, geographical and temporal coincidences, that the source of the Marseille-4 epidemic was the mink. The Marseille-4 variant was progressively replaced from January 2021 by the epidemic linked to the Alpha (UK) variant. Meanwhile, it circulated in 76 countries and accounted for 31,299 genomes in the GISAID database as of 30 June 2021, mostly obtained from European countries (in 29,906 cases (96%)) (Fig. S3).

During July and August 2020, two additional variants, Marseille-2 and Marseille 5, emerged in our geographical area, which became the second and third most prevalent variants from week 11 of 2020 after the Marseille-4 variant. The Marseille-2 variant (B.1.177) most likely emerged in Spain, where it accounted for two-thirds of released genomes and it spread to our geographical area and through Europe during the summer of 2020 (*19*). The Marseille-5 variant (B.1.367) disappeared in October 2020 and its source and origin are unknown. The Marseille-8 and Marseille-9 variants were first detected in August 2020. They accounted overall for approximatively 1.2% of the cases of SARS-CoV-2 infections in our geographical area, but nonetheless circulated until March and January 2021, respectively. The source and origin of these variants are unknown.

The Marseille-501 variant emerged in Marseille in January 2021 as the first variant of lineage 19B harbouring amino acid substitution N501Y in the spike protein (*25*). A majority of the genomes primarily available from the GISAID database were linked to Mayotte, which is part of the Comoros archipelago located in the Indian Ocean from where originated some of the patients we diagnosed to be infected with this variant. The Marseille-484K.V3-20A variant emerged in April 2021 in Marseille and was the majority variant apart from Beta (South African) and Gamma (Brazilian) variants to harbour amino acid substitution E484K in the spike protein. This variant was first described in Nigeria on 11 December 2020, but we have not documented its importation from this country.

The Alpha variant emerged in December 2020 in France and in Marseille and faded away in June 2021 after having been responsible for the majority of SARS-CoV-2 infections from February 2021. The first cases were part of a family cluster during the Christmas holidays after some family members came from England by train. We also had smaller epidemics with the Beta (South African) and the Gamma (Brazilian) variants from January 2021. Regarding the Gamma variant, we first detected it among people returning from the Comoros who had travelled through Ethiopia then Tunisia to Marseille. Between February and April 2021, we also detected three cases of a rare spike E484K-harbouring SARS-CoV-2, which we named Marseille-484K.V1 (*26*). The three genomes were phylogenetically closest to a genome from the Republic of Guinea (a.k.a. Guinea-Conakry), where one of the patients had travelled.

Most recently, we have mostly detected infections with the Delta (B.617.2) variant, in 4,113 patients. The two first patients diagnosed with this variant in late April 2021 were an Indian sailor re-embarking in Marseille after the resumption of maritime travel (La Scola *et al.*, 2021), and a physiotherapist who had travelled to the Republic of the Maldives, in Southeast Asia, and provided care upon his return to France in an accommodation facility for dependent elderly people, being involved in a cluster among residents and health care workers. Finally, in mid-July 2021, we detected a variant classified in the Pangolin lineage B.1.621, that was first identified in Colombia in January 2021. Information was available for one of the six patients infected with this variant, a 10-year-old Colombian girl who developed symptoms and was diagnosed in our institute four days after she arrived in France.

Overall, we identified several SARS-CoV-2 variants that were responsible as early as the summer of 2020 for distinct epidemics causing several waves of SARS-CoV-2 incidence (Figs. 6A, 6B, and 7) (*22, 23*). The epidemics caused by these different variants each exhibited a bell-shaped curve, and they occurred successively or concomitantly, therefore contributing to the total case load. We documented that new SARS-CoV-2 variants were introduced in our geographical area by boat in one case (Marseille-1), by train in one case (Alpha), and by plane in five cases (Alpha, Beta, Marseille-501, Marseille-484K.V1, B.1.617.V2, and B.1.621 variants). In addition, the importation of Marseille-484K.V3 from Nigeria through air travel is suspected. Importation from abroad through travel therefore accounted for a substantial source of variants and, subsequently, of cases. The other source identified for the emergence of a new variants is the mink epidemics in Northern France, with the case of Marseille-4 (*4*).

## DISCUSSION

In this study, we wanted to clarify the classification and naming of SARS-CoV-2 genotypes we implemented in our institute in order to have a simple identification of SARS-CoV-2 variants and to characterize their mutations, their source, and spread. This is essential to get a better understanding of SARS-CoV-2 epidemiology. Since the SARS-CoV-2 emergence in France, we have performed in our institute extensive surveillance, identification and monitoring of SARS-CoV-2 variants. Currently, the viral genotype has been determined for 44% of the patients diagnosed in our institute and the viral genome was obtained in approximatively one fifth of patients diagnosed with SARS-CoV-2. As early as the summer of 2020, we increased our surveillance of SARS-CoV-2 genotypes which allowed us to decipher several features of the current SARS-CoV-2 pandemic on a local scale. Before the summer of 2020, very few studies had pointed out an increase of SARS-CoV-2 genetic and amino acid diversity. Noticeably, Tomaszewski *et al.* had reported new pathways of mutational changes in SARS-CoV-2 proteomes (*27*).

First, it appears relatively clear that in countries which have been isolated, either geographically (islands that have closed their access) or politically (China and Korea), there has been only one major epidemic episode with a bell-shaped incidence curve. In Europe and the United States, where such a policy has not been implemented or has been made impossible by political circumstances, several epidemics have occurred successively or concurrently with different viral variants. Marseille, which is a very particular city due to its geographical location, has for centuries been at the forefront of imports of epidemics including of plague and cholera (*28*), and operated in an identical way during the SARS-CoV-2 pandemic. Thus, we observed that the first diagnosed SARS-CoV-2 infections were imported cases involving people who had stayed in northwest Italy and at the French/Italian border. This preceded a first epidemic phase during which international borders were closed. Their reopening in early summer 2020 was followed by a short epidemic. This was linked to a SARS-CoV-2 variant that we named Marseille-1, which was imported from North Africa by boat and originated from sub-Saharan Africa, and was probably not very transmissible, since this epidemic ended two months after its emergence and did not spread beyond the Marseille region. Another variant, which we named Marseille-2 (B.1.177), was introduced to our geographical area during the summer of 2020 from Spain, where it appears to have originated (*19*). Later, in 2021, the Alpha (B.1.1.7, or UK) variant was first imported in Marseille by French people returning from the UK. In addition, in the Marseille population which includes lots of people of Comorian origin, we observed the onset of small epidemics with the Beta (B.1.351, or South African) variant among people travelling by plane from the Comoros, and with the Marseille-501 (A.27) variant that involved people of Comorian origin including from Mayotte (*25*). Similarly, the reopening of sea cruises in the spring of 2020 was associated with an importation of the Delta (B.1.617.2, or Indian) variant by an Indian sailor who had travelled from India to Marseille (*29*), and a second case identified of this variant was in a traveller returning from the Republic of the Maldives located in the Indian Ocean. Two other SARS-CoV-2 variants were imported in 2021 from the Republic of Guinea and from Colombia. We therefore observed that the importation from abroad through travel accounted for a substantial source of variants and subsequently of infections, highlighting the role of globalisation in the spread of SARS-CoV-2 pandemic. Marseille is a city with many immigrant adults and children, and its geographical location and harbour make it very open to travellers. This explains the importation of infectious agents of various origins, which has been recognised in Marseille since antiquity (*28*). The considerable role of travel in the introduction and subsequent spread of new SARS-CoV-2 variants on the national or continental scales has also been reported in a recent phylogeographical study that analysed genomic, epidemiological, and mobility data (including ours) collected from ten European countries between January and October 2020 (*30*). This study found that the intensity of international travel predicted the spread of SARS-CoV-2 with the introduction of new viral variants, and it estimated that more than half of the lineages that were spreading in late summer 2020 had been newly introduced to countries since mid-June 2020. Such importations into countries of SARS-CoV-2 variants and their impact on the pattern of incidence of SARS-CoV-2 cases have also been reported in other studies (*31*). This sharply confirms what we described as early as at the beginning of September 2020 (*2*) with the emergence of seven new SARS-CoV-2 including one (Marseille-1) for which we had already found a source (*3*). Thus, we were the first to consider and highlight that the re-increase of SARS-CoV-2 incidence observed during the summer of 2020 in Marseille and in France may have resulted from the importation of new variants rather than from a rebound of previous viral genotypes (*2, 3*). This aligns with the fact that countries for which cross-border travel was the most limited geographically and/or politically by closing the borders experienced a single epidemic peak, whereas countries with high levels of international travel experienced several epidemic peaks and longer periods with a significant incidence of infections. Hence, these findings indicate that while local control measures such as lockdowns and curfews may limit viral spread, they are inefficient to avoid the introduction of new viral genotypes. The role of local lockdowns in controlling the viral spread has been controversial. This measure has been found to be irrelevant by some researchers (*32*). In Japan, lockdown was not associated with a reduction of SARS-CoV-2 incidence, whereas travel restrictions were (*33*).

Second, our findings indicate that the role of epizootics in dense animal herds has been overlooked in the generation of variants, their transmission to humans, and their contribution to the SARS-CoV-2 pandemic. Mink was revealed as a major source of SARS-CoV-2 diversity and a key element in the expansion of the human pandemic (*8, 9*). Data have accumulated indicating that the current pandemic, which is primarily a zoonosis, continued being a zoonosis with the massive involvement of minks, while other animals, including domestic animals such as cats and dogs, have also been affected (*9*). It has been demonstrated that minks were a source of new variants that fuelled the pandemic in Denmark (*8*), and SARS-CoV-2 spike protein gene variants harbouring amino acid substitutions N501T and G142D were also found to circulate in both humans and minks in the United States (*34*). In Marseille, we accumulated indications that the Marseille-4 variant originated from mink. This hypothesis took us a long time to confirm due to the very late release, in March 2021, of the viral sequence obtained from a mink farm in Eure-et-Loir in mid-November 2020. Interestingly, the detection of two epidemics, in Marseille and retrospectively in Mayenne (Northwest France), appeared to be unrelated as the absence of SARS-CoV-2 genotyping initially carried out in Mayenne and Eure-et-Loir prevented the demonstration that they were due to the same variant and that virus transmission could have occurred from farm minks located in a neighbouring department. Interestingly, the Marseille-4 variant was responsible for a large epidemic through Europe and was probably considerably overlooked as the global concern about SARS-CoV-2 variants only emerged several months after we described it.

These findings show that SARS-CoV-2 genomic surveillance and the definition of variants are valuable for a better assessment of SARS-CoV-2 epidemiology. In addition, they allow correlating viral genotypes with the clinical severity and outcome of infections. Indeed, several studies have reported different clinical pattern of infections according to the SARS-CoV-2 variant (*35, 36*). We previously compared the clinical symptoms and outcomes of patients infected with different variants we identified. Compared with viruses of the Nextstrain clade 20A that circulated between February and May 2020 in Marseille, patients infected with the Marseille-1 variant were less likely to report dyspnoea and rhinitis and to be hospitalised (*35*). In addition, patients infected with the Marseille-4 variant were more likely to exhibit fever and to be hospitalised than those infected with the Marseille-1 variant, and they were also more likely to exhibit fever than those infected with a virus of Nextstrain clade 20A. Also, patients infected with N501Y-harbouring SARS-CoV-2 variants (namely, Alpha (B.1.1.7), Beta (B.1.351) and Gamma (P.1) variants) were less likely to be hospitalised than those infected with the Marseille-1 variant and with viruses of Nextstrain clade 20A that circulated between February and May 2020 in Marseille (*35*). Furthermore, a key issue from the present study is highlighting the fluctuations in incidence of SARS-CoV-2 infections that we observed resulted from the succession or combination of distinct epidemics caused by different viral variants rather than from the rebound of persisting lineages, which we primarily described in September 2020 and was confirmed at larger scale (*30*). Since then, these genetic evolutionary and epidemiological patterns continued to be observed and the surveillance of SARS-CoV-2 variants, the role of travel and globalisation in their spread, and the turnover of major VOC has come to the frontline, particularly with the Alpha, Beta, Gamma, and Delta variants (*37, 38*).

Overall, previous findings have revealed that epidemic waves mostly observed in developed Western countries are, in fact, distinct epidemics due to different imported SARS-CoV-2 variants. Therefore, as the major identified sources for these SARS-CoV-2 variants is cross border travel and animal reservoirs, efficient border controls and surveillance of mustelid farms are critical in order to avoid the introduction of new SARS-CoV-2 variants and the occurrence of new epidemics.

## MATERIALS AND METHODS

### Epidemic curves for SARS-CoV-2-associated deaths in various countries

We used the cumulated daily mortality from the COVID-19 data repository operated by Johns Hopkins University (*39*) available at https://github.com/CSSEGISandData/COVID-19 (accessed on 25 June 2021), which covers 221 countries and locations from 22 April 2020 to 24 June 2021. Data mining of time series is a complex scientific field where the analysis process must be tailored to the characteristics of the series and the clustering objectives (*40, 41*). Our objective was to cluster the dynamic of epidemics at country scale as reflected by COVID-19 mortality, taking in account the time location of the different series peaks but not the country mortality incidences. A threshold of 500 deaths was used for countries to analyse their SARS-CoV-2-associated death curves. We transformed the cumulative death counts into smoothed (seven days centred moving average) daily death rates. The resulting series were then clustered using the Pearson distance after normalisation as L2-normed lock-step distance measure (*42*) and the Ascendant Hierarchical Classification algorithm with Ward aggregation. For determining the resulting clusters, we used a dynamic cluster detection method (*43*) with a minimum cluster size set at five countries. We then mapped the geographic distribution of cluster members. The data mining process was done using ‘R’(https://www.r-project.org/) with the packages ‘vegan’, ‘forecast’, ‘dynamictreecut’ and ‘rnaturalearth’.

### Study period and clinical samples

SARS-CoV-2 genotyping was performed from nasopharyngeal samples tested between 27 February 2020 and 18 August 2021 (18 months) at the IHU Méditerranée Infection institute (https://www.mediterranee-infection.com/). Specimens with a cycle threshold value (Ct) lower than 20 were selected as a in priority for whole genome sequencing of SARS-CoV-2 genomes, and those with a Ct between 20 and 30 were included secondarily to ensure a more comprehensive coverage of the study period. Partial sequencing of the SARS-CoV-2 spike gene generating a 1,854 nucleotide-long sequence was performed in 2020 and, mainly, between January and April 2021, as previously described (*25*). For respiratory specimens with Ct values > 30 or those with Ct values < 30 but from which genome sequences were not obtained, we identified those harbouring specific Marseille genotypes using qPCR targeting variant-specific regions, as previously described for the Marseille-1 variant (*3*) and the Marseille-4 variant (*4, 18*). Additional in house and commercial variant-specific qPCR assays were used for other variants. The study was approved by the ethics committee of the University Hospital Institute Méditerranée Infection under No. 2020-016-3.

### Genome sequencing

Viral RNA was extracted from 200 µL of nasopharyngeal swab fluid using the EZ1 Virus Mini Kit v2.0 on an EZ1 Advanced XL instrument (Qiagen, Courtaboeuf, France) or the KingFisher Flex system (Thermo Fisher Scientific, Waltham, MA, USA), following the manufacturer’s instructions. SARS-CoV-2 genome sequences were obtained by next-generation sequencing with various procedures since February 2020 until August 2021: (i) with the Illumina technology using the Nextera XT paired end strategy on MiSeq instruments (Illumina Inc., San Diego, CA, USA), as previously described (*3*); (ii) with the Illumina COVIDSeq protocol on a NovaSeq 6000 instrument (Illumina Inc.) since April 2021; or (iii) with Oxford Nanopore technology (ONT) on MinION or GridION instruments (Oxford Nanopore Technologies Ltd., Oxford, UK), as previously described (*3*). Next-generation sequencing with ONT was performed without or with (since March 2021) synthesized cDNA amplification using a multiplex PCR protocol with ARTIC nCoV-2019 V3 Panel primers purchased from Integrated DNA technologies (IDT, Coralville, IA, USA) according to the ARTIC procedure (https://artic.network/). After its extraction, viral RNA was reverse-transcribed using SuperScript IV (ThermoFisher Scientific) prior to cDNA second strand synthesis with Klenow Fragment DNA polymerase (New England Biolabs, Beverly, MA, USA) when performing NGS on the Illumina MiSeq instrument (Illumina Inc.) (*3*), LunaScript RT SuperMix kit (New England Biolabs) when performing NGS with the ONT, or according to the COVIDSeq protocol (Illumina Inc.) following the manufacturer’s recommandations. Generated cDNA was purified using Agencourt AMPure XP beads (Beckman Coulter, Villepinte, France) and quantified using Qubit 2.0 fluorometer (Invitrogen, Carlsbad, CA, USA), and fragment sizes were analysed on an Agilent Fragment analyser 5200 (Agilent Inc., Palo Alto, CA). Next-generation sequencing performed on the NovaSeq 6000 instrument followed the Illumina COVIDSeq protocol (Illumina Inc.), which included first strand cDNA synthesis from extracted viral RNA; cDNA amplification with two COVIDSeq primer pools; fragmentation and tagging of PCR amplicons with adapter sequences; clean up; PCR amplification (7 cycles) of tagmented amplicons; pool, clean up, quantification and normalization of libraries; and library sequencing on a NovaSeq 6000 sequencing system SP flow cell (Illumina Inc.).

### Genome sequence analyses

Genome consensus sequences were generated by mapping on the SARS-CoV-2 genome GenBank accession no. NC_045512.2 (Wuhan-Hu-1 isolate) with the CLC Genomics workbench v.7 (with the following thresholds: 0.8 for coverage and 0.9 for similarity) (https://digitalinsights.qiagen.com/), as previously described (*3*), or the Minimap2 software (*44*). A frequency of the majoritary nucleotide ≥70% and a nucleotide depth ≥10 (when sequence reads were generated on the NovaSeq Illumina instrument (Illumina Inc.)) or ≥5 (when sequences reads were generated on the MiSeq Illumina instrument) were used. Detection of mutations was performed using the freebayes tool (https://github.com/freebayes/freebayes) (*45*) with a mapping quality score of 20. SAMtools was used for soft clipping of Artic primers (https://artic.network/), and to remove PCR duplicates (*46*). Sequences described in the present study have been deposited on the GISAID sequence database (https://www.gisaid.org/) (***12***) and can be retrieved online using the GISAID online search tool with “Marseille” as keyword, then selecting sequence names containing “IHU” or “MEPHI”. In addition, they have been deposited on the IHU Marseille Infection website: https://www.mediterranee-infection.com/sequences-genomiques-sars-cov-2-completes-partielles-sequences-spike-protein-jusquen-mai-2021/.

Numbers of nucleotide changes in the SARS-CoV-2 genomes and of amino acid changes in the SARS-CoV-2 spike protein were obtained using the Nextclade tool (https://clades.nextstrain.org/results). They were plotted using the GraphPad software v5.01 (https://www.graphpad.com/). Pairwise nucleotide distances between SARS-CoV-2 genomes were computed using the MEGA7 software v10.2.5 (https://www.megasoftware.net/).

### Real-time reverse transcription-PCR specific of Marseille variants

For specimens with Ct values > 30 or those with Ct values < 30 but from which genome sequences were not obtained, we identified those harbouring specific Marseille genotypes using qPCR targeting variant-specific regions. We previously described qPCR specific of the Marseille-1 variant (*3*) and the Marseille-4 variant (*4*). We used other in house variant-specific qPCR systems for which the sequences of primers and probes are as follows, provided in 5’-3’ orientation. For the Beta (B.1.351) variant, qPCR targets the envelope gene and uses forward primer C_SA_3_MBF: TGAATTGCAGACACCTTTTGA, reverse primer C_SA_3_MBR: CAACCCTTGGTTGAATAGTCTTG, and probe C_SA_3_MBP: TGACATCTTCAATGGGGAATGT. For the Gamma (P.1) variant, qPCR targets the nucleocapsid gene and uses forward primer C_Bra_2_MBF: GTCAAGCCTCTTCTCGTTCCT, reverse primer C_Bra_2_MBR: AAAGCAAGAGCAGCATCACC, and probe C_Bra_2_MBP: GCAGCTCTAAACGAACTTCTCCTG. For the Delta (B.1.617.2) variant, two qPCR were used that target the envelope gene: the first one uses forward primer C_IND_1_MBF: AATCTTGATTCTAAGGTTGGTGGT, reverse primer C_IND_1_MBR: TGCTACCGGCCTGATAGATT, and probe C_IND_1_MBP: TTACCGGTATAGATTGTTTAGGAAGTCT; the second one uses forward primer P681_MBF: TGACATACCCATTGGTGCAG, reverse primer P681_MBR: GGCAATGATGGATTGACTAGC, and probe P681_MBP: ATTCTCATCGGCGGGCA. Finally, for the Marseille-2 (B.1.177) variant, qPCR targets the nsp16 gene and uses forward primer C2_2_MBF: CATGGTGGACAGCCTTTGTT, reverse primer C2_2_MBR: CATCTATTTGTTCGCGTGGTT, and probe C2_2_MBP: AATGCCTCATCATCTGAAGCAT. PCR conditions were the same than those described in (*25*), (*3*) and (*4*). Finally, we used the Applied Biosystems TaqPath COVID-19 kit (Thermo Fisher Scientific, Waltham, USA) then the TaqMan SARS-CoV-2 mutation assay H69-V70 (Thermo Fisher Scientific) for Alpha (B.1.1.7) variant detection, and TaqMan SARS-CoV-2 mutation assays L452R and P681R (Thermo Fisher Scientific) to detect the Delta (B.1.617.2) variant.

### SARS-CoV-2 classification and naming system

Phylogeny reconstructions based on the SARS-CoV-2 genomes obtained in our laboratory were performed using the nextstrain/ncov tool (https://github.com/nextstrain/ncov) then visualised with Auspice (https://docs.nextstrain.org/projects/auspice/en/stable/) or FigTree v1.4.4 (http://tree.bio.ed.ac.uk/software/figtree/); the limitation of sampling per date of collection of respiratory specimens was desactivated. Genome sequences were kept for analyses when larger than 24,000 nucleotides. Within reconstructed trees, we identified and counted all mutations between nodes and between nodes and leaves. This allowed us to define the set of mutations for the most recent ancestor of the clusters corresponding to each of the variants.

### Statistical analyses

Statistical tests were carried out using R 4.0.2 (https://cran.r-project.org/) and GraphPad software v5.01 (https://www.graphpad.com/). A p< 0.05 was considered statistically significant.

## Data Availability

SARS-CoV-2 genome sequences have been submitted to the GISAID sequence database (https://www.gisaid.org/).

## Acknowledgments

We are thankful to the IHU Méditerranée Infection COVID-19 Task Force that notably includes Sophie Amrane, Camille Aubry, Sofiane Bakour, Karim Bendamardji, Cyril Berenger, Nadim Cassir, Anne Darmon, Claire Decoster, Catherine Dhiver, Barbara Doudier, Sophie Edouard, Véronique Filosa, Séverine Guitton, Marie Hocquart, Morgane Mailhe, Florine Pelisson, Coralie Porcheto, Isabelle Ravaux, Piseth Seng, Laurence Thomas, Raphael Tola, Christelle Tomei, Catherine Triquet. This manuscript has been edited for English language by a native English speaker.

## Author contributions

Conceptualization, D.R., P.C., and P.-E.F.; methodology, D.R., P.C., P.E.F., A.L.; data and investigation, P.C., P.-E.F., H.C., J.D., A.G.-G., L.H., C.A., L.B., M.B., E.P., C.G., M.Be., E.B., P.D., H.T.-D., P.G., J.-C.L., M.M., P.B., P.P., F.F., M.D., B.L.S., A.L.; original draft preparation, D.R., P.C., and P.-E.F.; review and editing of the manuscript, all authors; supervision, D.R.;. All authors have read and agreed to the published version of the manuscript.

## Data Availability Statement

Genome sequences have been submitted to the GISAID sequence database (https://www.gisaid.org/).

## Ethics

All data were generated as part of the routine work at Assistance Publique-Hôpitaux de Marseille (Marseille university hospitals), and this study results from routine standard clinical management. The study was approved by the ethics committee of the University Hospital Institute Méditerranée Infection under No. 2020-016-3.

## REFERENCES

1. D. Cucinotta, M. Vanelli. WHO Declares COVID-19 a Pandemic. Acta Biomed. 91, 157–160 (2020).

2. P. Colson et al. Dramatic increase in the SARS-CoV-2 mutation rate and low mortality rate during the second epidemic in summer in Marseille. *IHU. pre-prints* (2020). https://doi.org/10.35088/68c3-ew82.

3. P. Colson et al. Introduction into the Marseille geographical area of a mild SARS-CoV-2 variant originating from sub-Saharan Africa: An investigational study. Travel. Med. Infect Dis 40, 101980 (2021).

4. P. E. Fournier et al. Emergence and outcomes of the SARS-CoV-2 ’Marseille-4’ variant. Int J Infect Dis 106, 228–236 (2021).

5. R. R. Sokal. Classification: purposes, principles, progress, prospects. Science. 185, 1115–1123 (1974).

6. E. Domingo, C. Perales. Viral quasispecies. PLoS. Genet. 15, e1008271 (2019).

7. M. Eigen. On the nature of virus quasispecies. Trends Microbiol. 4, 216–218 (1996).

8. B. B. Oude Munnink et al. Transmission of SARS-CoV-2 on mink farms between humans and mink and back to humans. Science 371, 172–177 (2021).

9. F. Fenollar et al. Mink, SARS-CoV-2, and the Human-Animal Interface. Front Microbiol. 12, 663815 (2021).

10. D. L. van Dorp et al. No evidence for increased transmissibility from recurrent mutations in SARS-CoV-2. Nat. Commun. 11, 5986–19818 (2020).

11. A. Rambaut et al. A dynamic nomenclature proposal for SARS-CoV-2 lineages to assist genomic epidemiology. Nat. Microbiol. 5, 1403–1407 (2020).

12. E. Alm et al. Geographical and temporal distribution of SARS-CoV-2 clades in the WHO European Region, January to June 2020. Euro. Surveill 25, 2001410 (2020).

13. J. Hadfield et al. Nextstrain: real-time tracking of pathogen evolution. Bioinformatics. 34, 4121–4123 (2018).

14. M. Chinazzi et al. The effect of travel restrictions on the spread of the 2019 novel coronavirus (COVID-19) outbreak. Science. 368, 395–400 (2020).

15. P. Colson et al. Ultrarapid diagnosis, microscope imaging, genome sequencing, and culture isolation of SARS-CoV-2. Eur. J Clin Microbiol Infect Dis. 39, 1601–1603 (2020).

16. A. Levasseur et al. Genomic diversity and evolution of coronavirus (SARS-CoV-2) in France from 309 COVID-19-infected patients. bioRxiv (2020). doi: https://doi.org/10.1101/2020.09.04.282616.

17. M. Bedotto, P. E. Fournier, L. Houhamdi, P. Colson, D. Raoult. Implementation of an in-house real-time reverse transcription-PCR assay to detect the emerging SARS-CoV-2 N501Y variants. J Clin. Virol 140, 104868 (2021).

18. M. Bedotto et al. Implementation of an in-house real-time reverse transcription-PCR assay for the rapid detection of the SARS-CoV-2 Marseille-4 variant. J Clin. Virol 139, 104814 (2021).

19. E. B. Hodcroft et al. Spread of a SARS-CoV-2 variant through Europe in the summer of 2020. Nature. 595, 707–712 (2021).

20. International Committee on Taxonomy of Viruses (ICTV). Virus Taxonomy: The ICTV Report on Virus Classification and Taxon Nomenclature. The Online (10th) Report of the International Committee on Taxonomy of Viruses. (2020). https://talk.ictvonline.org/ictv-reports/ictv_online_report/.

21. C. M. Fauquet et al. Geminivirus strain demarcation and nomenclature. Arch. Virol 153, 783–821 (2008).

22. Evolution of the weekly incidence of SARS-CoV-2 mutants and variants extrapolated to the total number of cases, based on their proportions of genotyped cases . Audiovisual Material (2021). https://doi.org/10.35081/nz2g-f980.

23. Introduction of SARS-CoV-2 mutants and variants in the Marseille geographical area through travel from abroad. Audiovisual Material (2021). https://doi.org/10.35081/ffd4-1y77.

24. Santé Publique France. Incidence rate of SARS-CoV-2 infections by French department per sliding week. (2021). https://geodes.santepubliquefrance.fr/.

25. P. Colson et al. Spreading of a new SARS-CoV-2 N501Y spike variant in a new lineage. Clin. Microbiol. Infect 27, 1352.e1-1352.e5. Online ahead of print. (2021).

26. P. Colson et al. Limited spread of a rare spike E484K-harboring SARS-CoV-2 in Marseille, France. IHU. pre-prints (2021). doi: https://doi.org/10.35088/2ngw-sa32.

27. T. Tomaszewski et al. New Pathways of Mutational Change in SARS-CoV-2 Proteomes Involve Regions of Intrinsic Disorder Important for Virus Replication and Release. Evol. Bioinform. Online. 16, 1176934320965149 (2020).

28. R. Barbieri, P. Colson, D. Raoult, M. Drancourt. Two-millennia fighting against port-imported epidemics, Marseille. I IHU. pre-prints (2021). doi: https://doi.org/10.35088/84a4-me41.

29. B. La Scola et al. SARS-CoV-2 variant from India to Marseille: the still active role of ports in the introduction of epidemics. Travel. Med Infect Dis 42, 102085 Epub 2021 May 21 (2021). doi: 10.1016/j.tmaid.2021.102085.

30. P. Lemey et al. Untangling introductions and persistence in COVID-19 resurgence in Europe. Nature. 595, 713–717 (2021).

31. A. McLaughlin et al. Early and ongoing importations of SARS-CoV-2 in Canada. medRxiv (2021). doi: https://doi.org/10.1101/2021.04.09.21255131.

32. E. R. Melnick, J. P. A. Ioannidis. Should governments continue lockdown to slow the spread of covid-19? BMJ.369 m1924 (2020). doi: 10.1136/bmj.m1924.

33. T. Hideki, M. Skidmore. A Cross-Country Analysis of the Determinants of Covid-19 Fatalities. CESifo Working Papers 9028 (2021). https://www.cesifo.org/en/publikationen/2021/working-paper/cross-country-analysis-determinants-covid-19-fatalities.

34. H. Y. Cai, A. Cai. SARS-CoV2 spike protein gene variants with N501T and G142D mutation-dominated infections in mink in the United States. J. Vet. Diagn. Invest. 33, 939–942 (2021).

35. T. L. Dao et al. Clinical outcomes in COVID-19 patients infected with different SARS-CoV-2 variants in Marseille, France. Clin. Microbiol. Infect. 10 (2021). S1198-743X(21)00270-6 Online ahead of print (2021). doi: 10.1016/j.cmi.2021.05.029.

36. P. Domingo et al. Not all COVID-19 pandemic waves are alike. Clin. Microbiol. Infect. 27, 1040–1040 (2021).

37. W. T. Harvey et al. SARS-CoV-2 variants, spike mutations and immune escape. Nat. Rev. Microbiol. 1–16 (2021).

38. R. C. Del, P. N. Malani, S. B. Omer. Confronting the Delta Variant of SARS-CoV-2, Summer 2021. JAMA. 10 (2021).

39. E. Dong, H. Du, L. Gardner. An interactive web-based dashboard to track COVID-19 in real time. Lancet Infect. Dis. 20, 533–534 (2020).

40. T. W. Liao. Clustering of time series data—a survey. Pattern Recognition 38, 1857–1874 (2005).

41. T. Fu. A review on time series data mining. Eng. Appl. Artif. Intell. 24, 164–181 (2011).

42. U. Mori, A. Mendiburu, J. A. Lozano. Similarity Measure Selection for Clustering Time Series Databases. IEEE Transactions on Knowledge and Data Engineering 28, 181–194 (2016).

43. P. Langfelder, B. Zhang, S. Horvath. Defining clusters from a hierarchical cluster tree: the Dynamic Tree Cut package for R. Bioinformatics. 24, 719–720 (2008).

44. H. Li. Minimap2: pairwise alignment for nucleotide sequences. Bioinformatics. 34, 3094–3100 (2018).

45. E. Garrison, G. Marth. Haplotype-based variant detection from short-read sequencing. *arXiv* (2012). https://arxiv.org/abs/1207.3907.

46. H. Li et al. The Sequence Alignment/Map format and SAMtools. Bioinformatics. 25, 2078–2079 (2009).

47. M. J. Adams, E. J. Lefkowitz, A. M. King, E. B. Carstens. Recently agreed changes to the International Code of Virus Classification and Nomenclature. Arch. Virol. 158, 2633–2639 (2013).

48. C. M. Fauquet, J. Stanley. Revising the way we conceive and name viruses below the species level: a review of geminivirus taxonomy calls for new standardized isolate descriptors. Arch. Virol 150, 2151–2179 (2005).

49. M. H. Van Regenmortel. Virus species and virus identification: past and current controversies. Infect Genet. Evol. 7, 133–144 (2007).

50. E. Domingo, R. A. Flavell, C. Weissmann. In vitro site-directed mutagenesis: generation and properties of an infectious extracistronic mutant of bacteriophage Qbeta. Gene 1, 3–25 (1976).

51. M. Eigen. Molecular self-organization and the early stages of evolution. Q. Rev. Biophys. 4, 149–212 (1971).

52. R. Andino, E. Domingo. Viral quasispecies. Virology 479-480, 46–51 (2015).

53. M. Eigen. Viral quasispecies. Sci. Am. 269, 42–49 (1993).

54. E. V. Koonin. Darwinian evolution in the light of genomics. Nucleic Acids Res. 37, 1011–1034 (2009).

55. A. Rambaut, D. Posada, K. A. Crandall, E. C. Holmes. The causes and consequences of HIV evolution. Nat. Rev. Genet. 5, 52–61 (2004).

56. J. H. Kuhn et al. Virus nomenclature below the species level: a standardized nomenclature for laboratory animal-adapted strains and variants of viruses assigned to the family Filoviridae. Arch. Virol. 158, 1425–1432 (2013).

