## Supplementary Material for "Analysis of SARS-CoV-2 variants from 24,181 patients exemplifies the role of globalisation and zoonosis in pandemics"

**Supplementary Figures**

**Fig. S1. Chronological distribution of SARS-CoV-2 diagnoses by qPCR at IHU Méditerranée Infection institute (A) and mean number per month of amino acid substitutions in the SARS-CoV-2 spike (B).**

(**B**) Number per month of amino acid substitutions in SARS-CoV-2 spike proteins are calculated in reference to the genome of the Wuhan-Hu-1 isolate (GenBank Accession no. NC_045512.2).

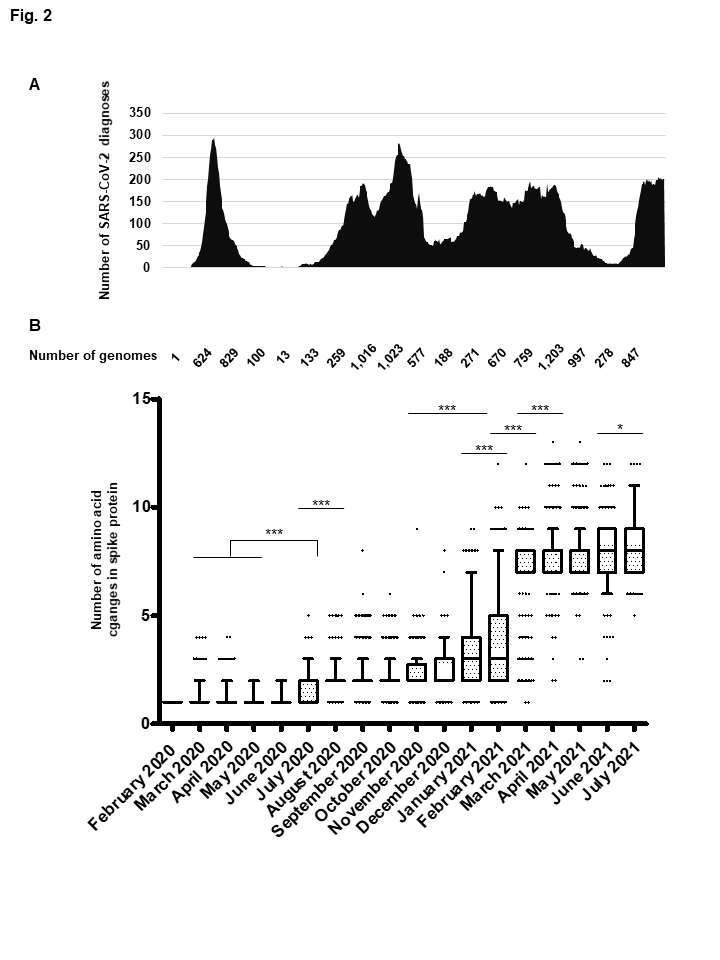

**Fig. S2. Timeline of trends of searches in Google for world (a) and France (b) and number of publications (c, d) with “variant” and “Covid-19” or “SARS-CoV-2” as keywords.**

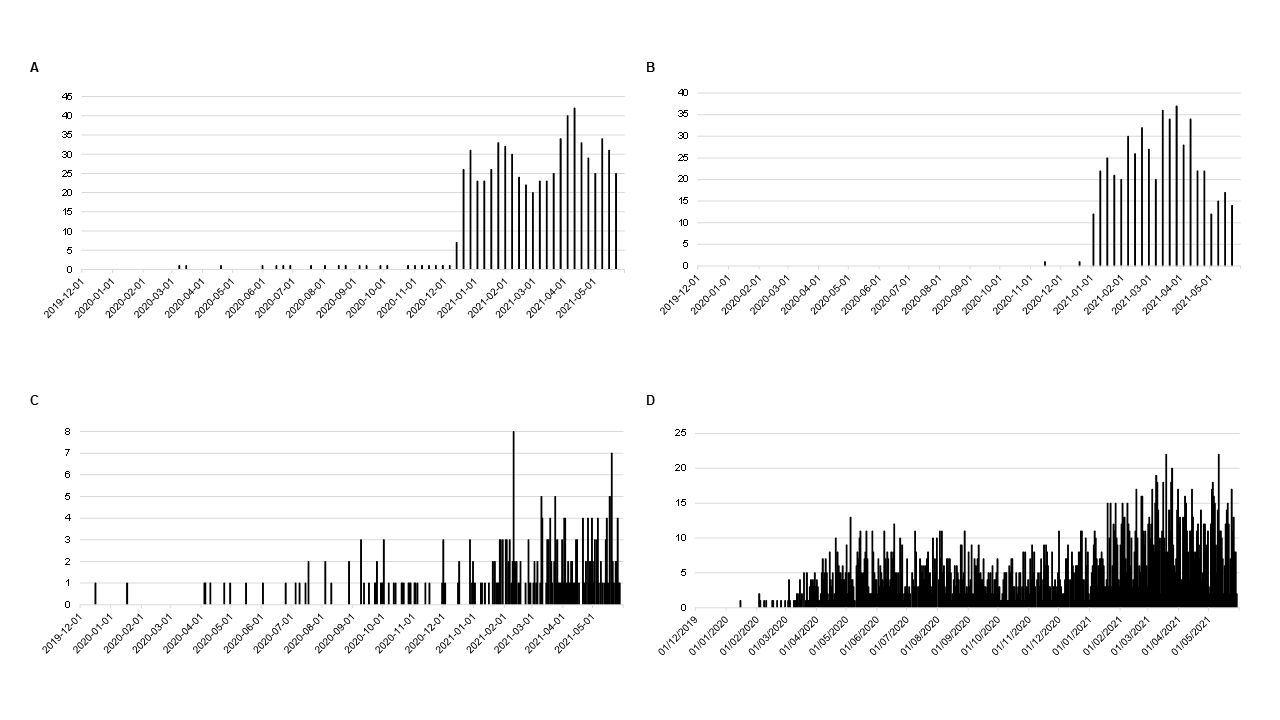

**Fig. S3. The emergence and outcome of the SARS-CoV-2 Marseille-4 variant (according to GISAID database as of 30 June 2021).**

Number of genomes per day (A), number of genomes per day and country (B), and time range of genome collection and total number of genomes per world region (C) are shown.

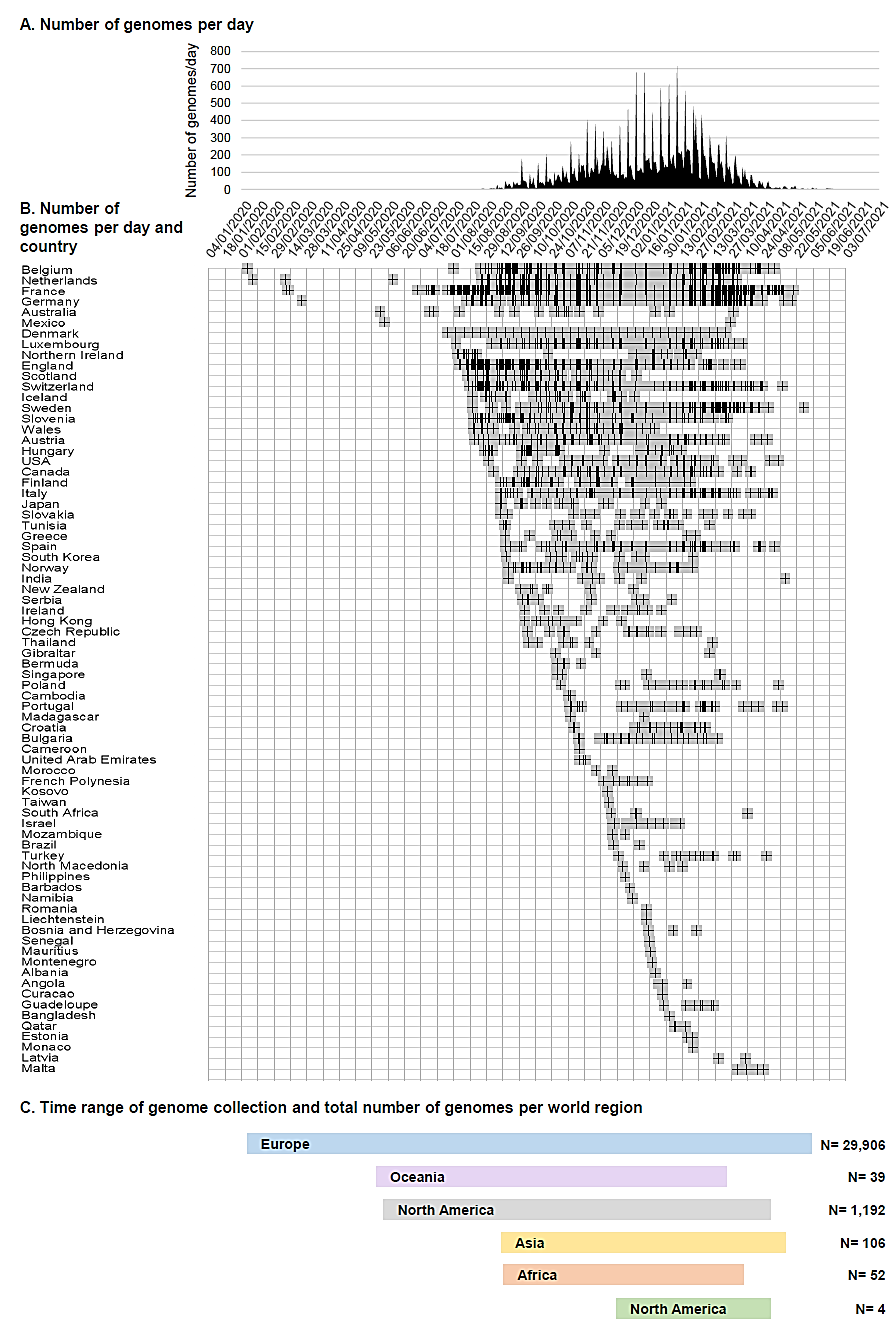

**Supplementary Tables**

**Supplementary Table S1.** **Monthly numbers of genome sequences, spike sequences and qPCR performed each month, and proportions of positive diagnoses of SARS-CoV-2 infection as of July 2021.**

| **Month/Year** | **Total number of positive diagnoses** |  | **SARS-CoV-2 genotyping approach** | | | | | | | |
| --- | --- | --- | --- | --- | --- | --- | --- | --- | --- | --- |
|  |  |  | **Next-generation genome sequencing** | |  | **Next-generation spike gene fragment sequencing** | |  | **qPCR** | |
|  |  |  | **Number of genomes** | ***% of the positive diagnoses*** |  | **Number of sequences** | ***% of the positive diagnoses*** |  | **Number of positive** | ***% of the positive diagnoses*** |
| 2020/2 | 6 |  | 3 | *50* |  | 1 | *17* |  | 0 | *0,0* |
| 2020/3 | 3,731 |  | 777 | *21* |  | 1 | *<0,1* |  | 7 | *0,2* |
| 2020/4 | 2,790 |  | 877 | *31* |  | 2 | *0,1* |  | 5 | *0,2* |
| 2020/5 | 207 |  | 70 | *34* |  | 0 | *0,0* |  | 1 | *0,5* |
| 2020/6 | 48 |  | 23 | *48* |  | 0 | *0,0* |  | 0 | *0,0* |
| 2020/7 | 368 |  | 244 | *66* |  | 0 | *0,0* |  | 2 | *0,5* |
| 2020/8 | 2,620 |  | 741 | *28* |  | 1 | *<0,1* |  | 15 | *0,6* |
| 2020/9 | 4,664 |  | 1,192 | *25* |  | 1 | *<0,1* |  | 16 | *0,3* |
| 2020/10 | 6,413 |  | 1,713 | *27* |  | 0 | *0,0* |  | 19 | *0,3* |
| 2020/11 | 3,911 |  | 751 | *19* |  | 1 | *<0,1* |  | 26 | *0,7* |
| 2020/12 | 2,116 |  | 450 | *21* |  | 1 | *0,1* |  | 213 | *10* |
| 2021/1 | 4,391 |  | 692 | *16* |  | 50 | *1* |  | 2,445 | *56* |
| 2021/2 | 5,928 |  | 1,075 | *18* |  | 992 | *17* |  | 2,751 | *46* |
| 2021/3 | 6,016 |  | 724 | *12* |  | 2,639 | *39* |  | 1,879 | *31* |
| 2021/4 | 6,210 |  | 1,503 | *24* |  | 0 | *0,0* |  | 3,667 | *59* |
| 2021/5 | 2,566 |  | 1,137 | *44* |  | 0 | *0,0* |  | 1,199 | *47* |
| 2021/6 | 660 |  | 278 | *42* |  | 0 | *0,0* |  | 293 | *44* |
| 2021/7 | 3,767 |  | 1,412 | *37* |  | 0 | *0,0* |  | 1,674 | *44* |

**Supplementary Table S2. Mean (±standard deviation) numbers per month of mutations in SARS-CoV-2 genomes obtained from patients SARS-CoV-2-diagnosed at IHU Méditerranée Infection, Marseille.**

Numbers of mutations in SARS-CoV-2 genomes are calculated in reference to the genome of the Wuhan-Hu-1 isolate (GenBank Accession no. NC_045512.2). Whiskers indicate 10-90 percentiles.

|  | February 2020 | March 2020 | April 2020 | May 2020 | June 2020 | July 2020 | August 2020 | September 2020 | October 2020 | November 2020 | December 2020 | January 2021 | February 2021 | March 2021 | April 2021 | May 2021 | June 2021 | July 2021 |
| --- | --- | --- | --- | --- | --- | --- | --- | --- | --- | --- | --- | --- | --- | --- | --- | --- | --- | --- |
| **Number of genomes analyzed** | 1 | 624 | 829 | 100 | 13 | 133 | 259 | 1,016 | 1,023 | 577 | 188 | 271 | 670 | 759 | 1,203 | 997 | 278 | 847 |
| **Nucleotide changes in the genome** |  |  |  |  |  |  |  |  |  |  |  |  |  |  |  |  |  |  |
| Median | 5 | 8 | 8 | 9 | 11 | 13 | 21 | 21 | 22 | 24 | 26 | 24 | 27 | 34 | 36 | 36 | 37 | 36 |
| Mean | 5.0 | 8.1 | 8.5 | 9.4 | 10.8 | 15.0 | 20.0 | 20.5 | 21.9 | 23.0 | 25.2 | 24.2 | 26.4 | 34.1 | 35.7 | 36.2 | 36.8 | 36.1 |
| Std. Deviation | 0.0 | 2.0 | 2.0 | 2.3 | 2.7 | 4.4 | 5.0 | 4.7 | 4.1 | 4.1 | 4.3 | 5.2 | 5.7 | 4.2 | 3.8 | 3.9 | 4.7 | 3.1 |
| **Amino acid substitutions in the spike** | |  |  |  |  |  |  |  |  |  |  |  |  |  |  |  |  |  |
| Median | 1 | 1 | 1 | 1 | 1 | 2 | 2 | 2 | 2 | 2 | 2 | 3 | 3 | 7 | 7 | 8 | 8 | 8 |
| Mean | 1.0 | 1.1 | 1.1 | 1.1 | 1.2 | 1.7 | 2.2 | 2.2 | 2.2 | 2.3 | 2.5 | 3.3 | 3.8 | 7.1 | 7.7 | 7.8 | 7.8 | 8.2 |
| Std. Deviation | 0.0 | 0.5 | 0.4 | 0.3 | 0.4 | 0.8 | 0.7 | 0.7 | 0.7 | 0.7 | 1.0 | 2.0 | 2.2 | 1.3 | 1.0 | 1.1 | 1.5 | 1.5 |

**Supplementary Table S3. Weekly proportions of the variants among patients SARS-CoV-2-diagnosed in our institute.**

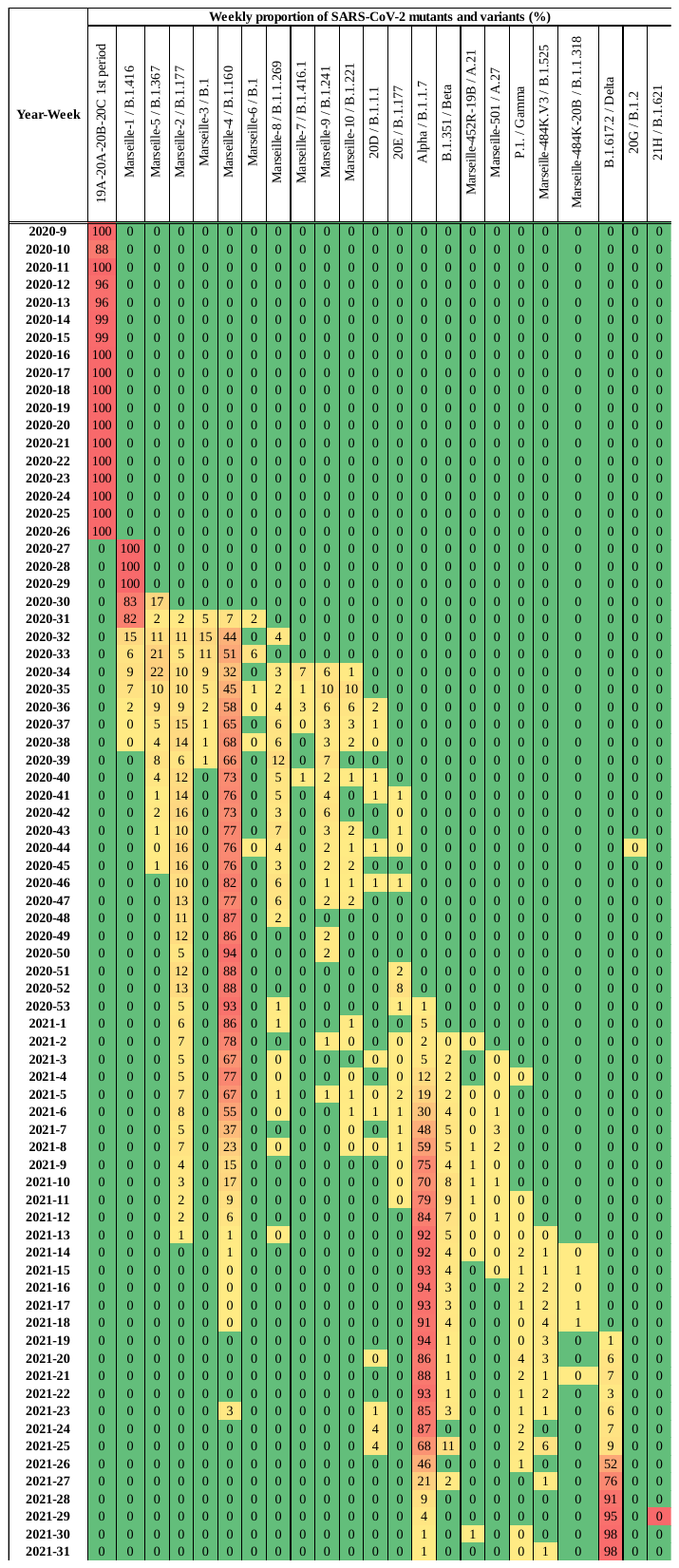
